# REACT-1 round 13 final report: exponential growth, high prevalence of SARS-CoV-2 and vaccine effectiveness associated with Delta variant in England during May to July 2021

**DOI:** 10.1101/2021.09.02.21262979

**Authors:** Paul Elliott, David Haw, Haowei Wang, Oliver Eales, Caroline E. Walters, Kylie E. C. Ainslie, Christina Atchison, Claudio Fronterre, Peter J. Diggle, Andrew J. Page, Alexander J. Trotter, Sophie J. Prosolek, The COVID-19 Genomics UK (COG-UK) Consortium, Deborah Ashby, Christl A. Donnelly, Wendy Barclay, Graham Taylor, Graham Cooke, Helen Ward, Ara Darzi, Steven Riley

**Author notes:** Corresponding authors: Paul Elliott and Steven Riley, School of Public Health, Imperial College London, Norfolk Place, London, W2 1PG. https://www.cogconsortium.uk (The COVID-19 Genomics UK (COG-UK) Consortium).

## Abstract

**Background:** The prevalence of SARS-CoV-2 infection continues to drive rates of illness and hospitalisations despite high levels of vaccination, with the proportion of cases caused by the Delta lineage increasing in many populations. As vaccination programs roll out globally and social distancing is relaxed, future SARS-CoV-2 trends are uncertain.

**Methods:** We analysed prevalence trends and their drivers using reverse transcription-polymerase chain reaction (RT-PCR) swab-positivity data from round 12 (between 20 May and 7 June 2021) and round 13 (between 24 June and 12 July 2021) of the REal-time Assessment of Community Transmission-1 (REACT-1) study, with swabs sent to non-overlapping random samples of the population ages 5 years and over in England.

**Results:** We observed sustained exponential growth with an average doubling time in round 13 of 25 days (lower Credible Interval of 15 days) and an increase in average prevalence from 0.15% (0.12%, 0.18%) in round 12 to 0.63% (0.57%, 0.18%) in round 13. The rapid growth across and within rounds appears to have been driven by complete replacement of Alpha variant by Delta, and by the high prevalence in younger less-vaccinated age groups, with a nine-fold increase between rounds 12 and 13 among those aged 13 to 17 years. Prevalence among those who reported being unvaccinated was three-fold higher than those who reported being fully vaccinated. However, in round 13, 44% of infections occurred in fully vaccinated individuals, reflecting imperfect vaccine effectiveness against infection despite high overall levels of vaccination. Using self-reported vaccination status, we estimated adjusted vaccine effectiveness against infection in round 13 of 49% (22%, 67%) among participants aged 18 to 64 years, which rose to 58% (33%, 73%) when considering only strong positives (Cycle threshold [Ct] values < 27); also, we estimated adjusted vaccine effectiveness against symptomatic infection of 59% (23%, 78%), with any one of three common COVID-19 symptoms reported in the month prior to swabbing. Sex (round 13 only), ethnicity, household size and local levels of deprivation jointly contributed to the risk of higher prevalence of swab-positivity.

**Discussion:** From end May to beginning July 2021 in England, where there has been a highly successful vaccination campaign with high vaccine uptake, infections were increasing exponentially driven by the Delta variant and high infection prevalence among younger, unvaccinated individuals despite double vaccination continuing to effectively reduce transmission. Although slower growth or declining prevalence may be observed during the summer in the northern hemisphere, increased mixing during the autumn in the presence of the Delta variant may lead to renewed growth, even at high levels of vaccination.

## Introduction

Despite the successful development, licensing and distribution of effective vaccines against COVID-19 [1,2], the number of newly reported cases and deaths continued to rise globally into the northern hemisphere summer of 2021 [3]. Prior trends of decreasing prevalence were being reversed in some populations where the Delta variant had become dominant, leading to estimates of a substantially higher transmissibility for Delta compared to Alpha [4]. In addition, globally, only 13% of the population are fully vaccinated while only 1% of people in low income countries have received even one dose [5]. Despite the potential for reduced growth during the northern hemisphere summer, many countries are evaluating the possibility of a further large wave of infections in the autumn, driven by the Delta variant.

The incidence of reverse transcription-polymerase chain reaction (RT-PCR) confirmed cases of COVID-19 has increased substantially in England since the Delta variant became established during April to May 2021 [6]. Over the same period, the UK government proceeded with its gradual relaxation of social distancing (roadmap) [7] with the ending of almost all legal restrictions in England on 19 July 2021 [8]. While a much lower proportion of COVID-19 cases resulted in hospitalisations in England versus a comparable period of growth during autumn 2020, exponential growth in hospitalisations was still observed from mid-June 2021 [6].

Here, we describe the underlying dynamics driving patterns in SARS-CoV-2 infections from the end of May to the beginning of July 2021 in England by analysing RT-PCR swab-positivity data from the two most recent rounds of the REal-time Assessment of Community Transmission-1 (REACT-1) study [9,10].

## Results

Valid RT-PCR results were obtained from 108,911 participants in round 12 (20 May to 7 June 2021) and 98,233 participants in round 13 (24 June to 12 July 2021) (Table 1), recruited as non-overlapping random samples of the population aged 5 years and above (Methods).

**Table 1.**
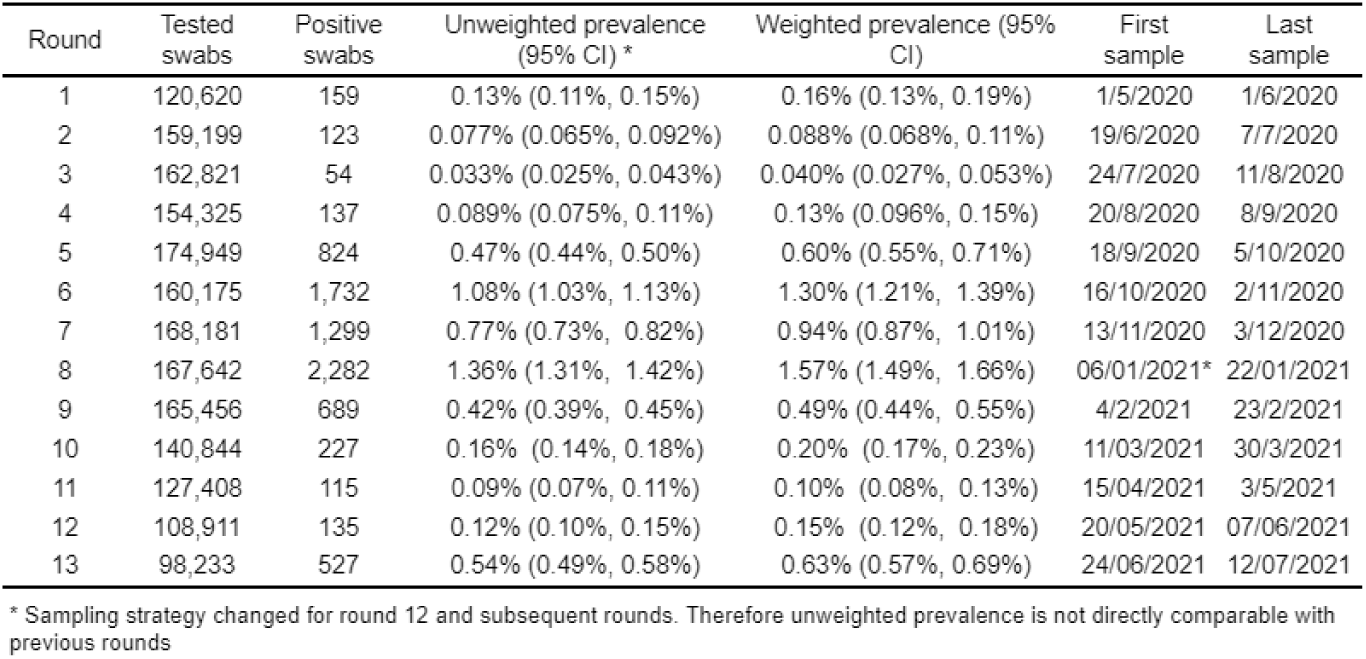
Unweighted and weighted prevalence of swab-positivity across 13 rounds of REACT-1.

### Prevalence and growth

Prevalence of infection with SARS-CoV-2 increased substantially in England between rounds 12 and 13 (Figure 1) as the third wave took hold, linked to the rapid replacement of Alpha by Delta variant. In round 13, between 24 June and 12 July 2021, we found 527 positives from 98,233 swabs giving a weighted prevalence of 0.63% (0.57%, 0.69%), and, on average, a greater than four-fold rise compared with the weighted prevalence in round 12 of 0.15% (0.12%, 0.18%) (Table 1). The prevalence in round 13 was similar to that observed in early October 2020 and late January 2021 during, respectively, the rise and fall of the second wave (Figure 1).

**Figure 1.**
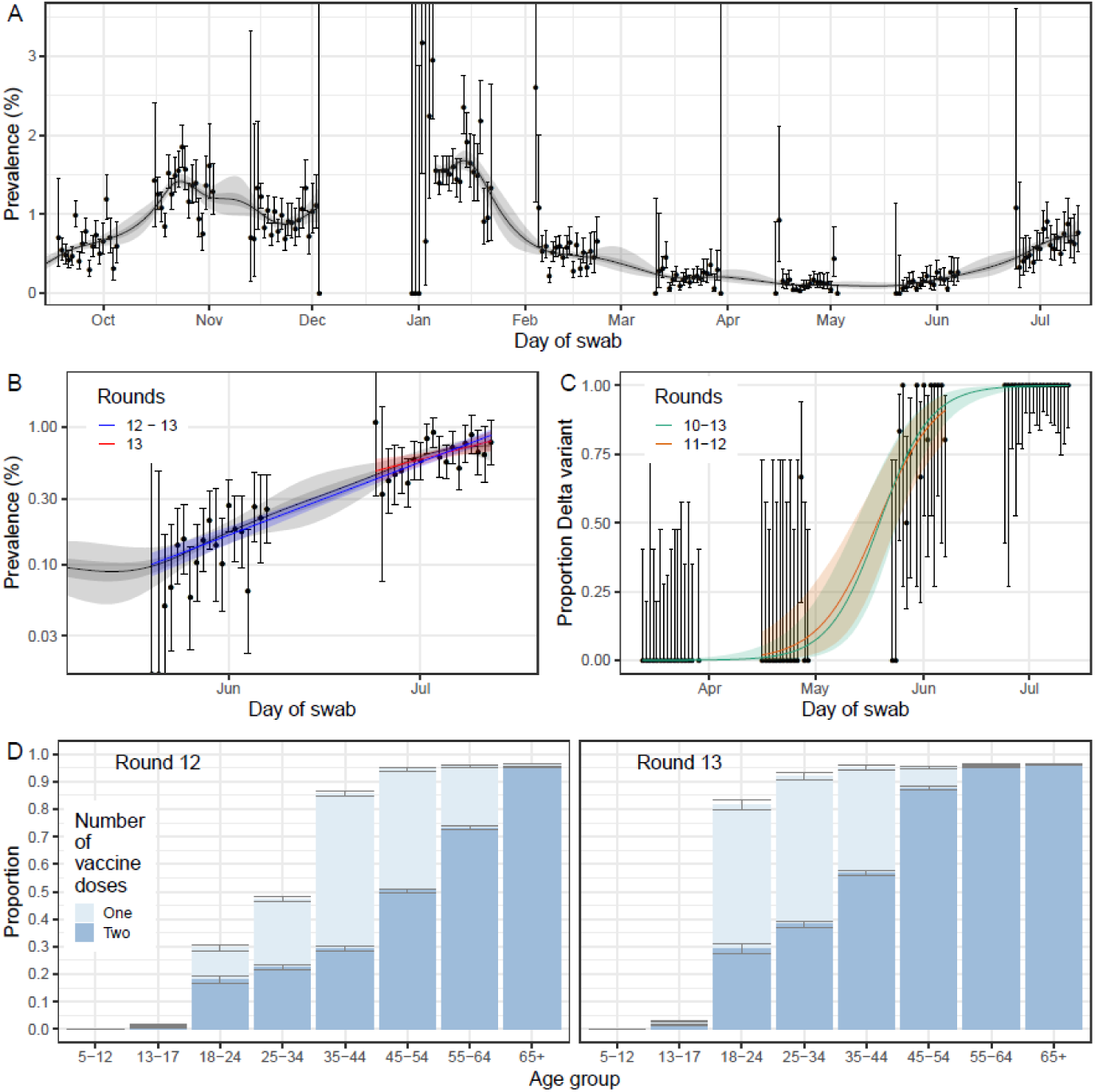
Temporal trends in prevalence, proportion of positive cases determined to be the Delta variant and vaccine coverage (A) Prevalence of national swab-positivity for England estimated using a P-spline for all thirteen rounds with central 50% (dark grey) and 95% (light grey) posterior credible intervals. Shown here from round 5 onwards of the study weighted observations (black dots) and 95% binomial confidence intervals (vertical lines) are also shown. Note that the period between round 7 and round 8 (December) of the model is not included as there were no data available to capture the late December peak of the epidemic.(B) Comparison of the exponential model fit to round 12 and 13 (blue) and the exponential model fit to round 13 only (red). Also shown is the P-spline model fit from panel A. Shown here only for rounds 12 and 13 of the study with a log_10_ y-axis. (C) Proportion of Delta against Alpha over time. Points show raw data with error bars representing the 95% confidence interval. Shaded regions show best fit Bayesian logistic regression models, fit to rounds 10 to 13 (green) and rounds 11 to 12 (orange), with 95% credible interval. (D) Proportion of individuals, for whom vaccine status is known, who reported being vaccinated with one (light blue) or two (dark blue) doses.

The Delta variant completely replaced Alpha during the period of our study. Of the 254 lineages determined for round 13, 100% were the Delta variant, compared with round 12 during which 36 of 46 (78.3%) were Delta and the remaining 10 were Alpha variant. Growth of Delta against Alpha for round 10 (11 to 30 March 2021) to round 13 corresponded to a daily growth rate advantage of 0.14 (0.10, 0.20) for Delta, which, in turn, corresponded to an additive R advantage of 0.86 (0.63, 1.23) (Figure 1). Within the Delta variant, we did not detect the K417N mutation associated with the AY.1 and AY.2 lineages. Under the assumption that REACT-1 participants provide an unbiased sample of infections, we can exclude, with 95% confidence, a population prevalence of non-Delta lineages greater than 0.004%, corresponding to 2,350 infections in England on average during round 13.

Nationally, we observed an exponential trend in prevalence with sustained growth for rounds 12 to 13 (between 20 May and 12 July 2021) (Table 2, Figure 1) despite England having one of the highest adult vaccination rates internationally [5]. Averaging over the period of each of rounds 12 and 13 separately, we estimated the reproduction number R at 1.44 (1.20, 1.73) (round 12) and 1.19 (1.06, 1.32) (round 13), corresponding to doubling times of 11 (7, 23) days and 25 days (with a lower Credible Interval [CI] of 15 days) respectively. Across rounds 12 to 13, R was 1.28 (1.24, 1.31) with a doubling time of 17 (15, 19) days. Patterns of growth for the period of the study were robust when considering alternative definitions of positivity, such as only non-symptomatic individuals or positive samples with lower cycle threshold (Ct) values, corresponding to higher viral load (Table 2).

**Table 2.** Estimates of national growth rates, doubling times and reproduction numbers for round 13, round 12 to round 13, and round 12.

| Round | Outcome | Growth rate | R | Probability<br>R>1 | Doubling (+) / Halving (-)<br>time |
| --- | --- | --- | --- | --- | --- |
| 13 | All positives | 0.028 ( 0.009 , 0.046 ) | 1.19 ( 1.06 , 1.32 ) | >0.99 | 25.0 ( * , 15.1 ) |
|  | Non-symptomatics | 0.042 ( 0.011 , 0.072 ) | 1.28 ( 1.07 , 1.52 ) | >0.99 | 16.7 ( * , 9.6 ) |
|  | Positive for both<br>E and N genes | 0.019 ( -0.002 , 0.041 ) | 1.13 ( 0.99 , 1.28 ) | 0.96 | 35.7 ( * , 16.9 ) |
|  | Positive for both<br>E and N genes or<br>positive only for N<br>gene with CT 35 or<br>less | 0.030 ( 0.011 , 0.050 ) | 1.20 ( 1.07 , 1.35 ) | >0.99 | 23.2 ( * , 13.9 ) |
| 12 and 13 | All positives | 0.041 ( 0.036 , 0.046 ) | 1.28 ( 1.24 , 1.31 ) | >0.99 | 17.0 ( 19.2 , 15.2 ) |
|  | Non-symptomatics | 0.042 ( 0.034 , 0.050 ) | 1.29 ( 1.23 , 1.35 ) | >0.99 | 16.6 ( 20.5 , 13.8 ) |
|  | Positive for both<br>E and N genes | 0.042 ( 0.036 , 0.048 ) | 1.29 ( 1.25 , 1.33 ) | >0.99 | 16.5 ( 19.1 , 14.5 ) |
|  | Positive for both<br>E and N genes or<br>positive only for N<br>gene with CT 35 or<br>less | 0.041 ( 0.036 , 0.046 ) | 1.28 ( 1.24 , 1.32 ) | >0.99 | 17.0 ( 19.4 , 15.1 ) |
| 12 | All positives | 0.063 ( 0.030 , 0.097 ) | 1.44 ( 1.20 , 1.73 ) | >0.99 | 11.1 ( 23.4 , 7.1 ) |
|  | Non-symptomatics | 0.030 ( -0.027 , 0.087 ) | 1.20 ( 0.84 , 1.64 ) | 0.85 | 23.3 ( -26.0 , 8.0 ) |
|  | Positive for both<br>E and N genes | 0.104 ( 0.064 , 0.146 ) | 1.79 ( 1.45 , 2.18 ) | >0.99 | 6.7 ( 10.8 , 4.8 ) |
|  | Positive for both<br>E and N genes or<br>positive only for N<br>gene with CT 35 or<br>less | 0.073 ( 0.037 , 0.110 ) | 1.53 ( 1.25 , 1.84 ) | >0.99 | 9.5 ( 18.7 , 6.3 ) |

\* Doubling/Halving time had an estimated magnitude greater than 50 days and so represented approximately constant prevalence

Alongside the rapid rise of the Delta variant, recent growth in England appears to have been driven by younger age groups (Table 3, Figure 2). For example, weighted prevalence in round 13 was nine-fold higher in 13-17 year olds at 1.56% (1.25%, 1.95%) compared with 0.16% (0.08%, 0.31%) in round 12. Similar patterns were observed in England for the same period in a longitudinal household study [11]. More generally, participants aged between 5 and 24 years were over-represented among infected people in our study, contributing 50% of infections (weighted age-standardised) while only representing 25% of the population of England aged 5 years or above [12]. Therefore, during this period of rapid growth, any interventions targeted at the younger ages would have a disproportionate impact in slowing the epidemic [13].

**Table 3a.** Unweighted and weighted prevalence of swab-positivity for sex, age, and region for round 12 and round 13.

| Variable | Category | Round 12 |  |  |  | Round 13 |  |  |  |
| --- | --- | --- | --- | --- | --- | --- | --- | --- | --- |
|  |  | Positive | Total | Unweighted Prevalence | Weighted Prevalence* | Positive | Total | Unweighted Prevalence | Weighted Prevalence* |
| Gender | Male | 55 | 48,190 | 0.11% ( 0.09% , 0.15% ) | 0.14% ( 0.10% , 0.18% ) | 252 | 43,272 | 0.58% ( 0.51% , 0.66% ) | 0.71% ( 0.62% , 0.82% ) |
|  | Female | 80 | 60,718 | 0.13% ( 0.10% , 0.16% ) | 0.16% ( 0.12% , 0.20% ) | 275 | 54,957 | 0.50% ( 0.44% , 0.56% ) | 0.55% ( 0.48% , 0.63% ) |
|  | Unknown | 0 | 3 | 0.00% ( 0.00% , 70.76% ) | NA ( NA , NA ) | 0 | 4 | 0.00% ( 0.00% , 60.24% ) | 0 ( NA , NA ) |
| Age | 05-12 | 26 | 7,598 | 0.34% ( 0.22% , 0.50% ) | 0.35% ( 0.23% , 0.54% ) | 70 | 6,994 | 1.00% ( 0.78% , 1.26% ) | 1.02% ( 0.80% , 1.31% ) |
|  | 13-17 | 10 | 5,906 | 0.17% ( 0.08% , 0.31% ) | 0.16% ( 0.08% , 0.31% ) | 86 | 5,423 | 1.59% ( 1.27% , 1.95% ) | 1.56% ( 1.25% , 1.95% ) |
|  | 18-24 | 12 | 4,044 | 0.30% ( 0.15% , 0.52% ) | 0.36% ( 0.20% , 0.64% ) | 47 | 3,117 | 1.51% ( 1.11% , 2.00% ) | 1.56% ( 1.15% , 2.13% ) |
|  | 25-34 | 11 | 9,059 | 0.12% ( 0.06% , 0.22% ) | 0.11% ( 0.06% , 0.21% ) | 54 | 7,888 | 0.68% ( 0.51% , 0.89% ) | 0.72% ( 0.54% , 0.97% ) |
|  | 35-44 | 16 | 12,592 | 0.13% ( 0.07% , 0.21% ) | 0.12% ( 0.07% , 0.21% ) | 73 | 11,876 | 0.61% ( 0.48% , 0.77% ) | 0.61% ( 0.48% , 0.78% ) |
|  | 45-54 | 23 | 17,564 | 0.13% ( 0.08% , 0.20% ) | 0.14% ( 0.09% , 0.22% ) | 74 | 15,776 | 0.47% ( 0.37% , 0.59% ) | 0.46% ( 0.36% , 0.58% ) |
|  | 55-64 | 16 | 21,156 | 0.08% ( 0.04% , 0.12% ) | 0.08% ( 0.05% , 0.14% ) | 59 | 18,800 | 0.31% ( 0.24% , 0.40% ) | 0.31% ( 0.24% , 0.40% ) |
|  | 65-74 | 15 | 20,647 | 0.07% ( 0.04% , 0.12% ) | 0.07% ( 0.04% , 0.12% ) | 48 | 18,836 | 0.25% ( 0.19% , 0.34% ) | 0.25% ( 0.19% , 0.34% ) |
|  | 75+ | 6 | 10,345 | 0.06% ( 0.02% , 0.13% ) | 0.07% ( 0.03% , 0.18% ) | 16 | 9,523 | 0.17% ( 0.10% , 0.27% ) | 0.17% ( 0.10% , 0.29% ) |
| Region | South East | 21 | 18,855 | 0.11% ( 0.07% , 0.17% ) | 0.14% ( 0.09% , 0.23% ) | 61 | 17,427 | 0.35% ( 0.27% , 0.45% ) | 0.39% ( 0.29% , 0.51% ) |
|  | North East | 6 | 5,018 | 0.12% ( 0.04% , 0.26% ) | 0.14% ( 0.05% , 0.34% ) | 27 | 4,500 | 0.60% ( 0.40% , 0.87% ) | 0.74% ( 0.48% , 1.14% ) |
|  | North West | 22 | 13,229 | 0.17% ( 0.10% , 0.25% ) | 0.26% ( 0.16% , 0.41% ) | 81 | 11,814 | 0.69% ( 0.54% , 0.85% ) | 0.77% ( 0.59% , 0.99% ) |
|  | Yorkshire and The Humber | 16 | 10,538 | 0.15% ( 0.09% , 0.25% ) | 0.17% ( 0.10% , 0.29% ) | 70 | 9,550 | 0.73% ( 0.57% , 0.93% ) | 0.88% ( 0.67% , 1.15% ) |
|  | East Midlands | 12 | 9,145 | 0.13% ( 0.07% , 0.23% ) | 0.19% ( 0.10% , 0.36% ) | 47 | 8,425 | 0.56% ( 0.41% , 0.74% ) | 0.65% ( 0.47% , 0.89% ) |
|  | West Midlands | 12 | 10,993 | 0.11% ( 0.06% , 0.19% ) | 0.11% ( 0.06% , 0.20% ) | 49 | 9,842 | 0.50% ( 0.37% , 0.66% ) | 0.56% ( 0.40% , 0.77% ) |
|  | East of England | 17 | 12,388 | 0.14% ( 0.08% , 0.22% ) | 0.14% ( 0.08% , 0.24% ) | 42 | 11,210 | 0.37% ( 0.27% , 0.51% ) | 0.41% ( 0.30% , 0.58% ) |
|  | London | 22 | 16,966 | 0.13% ( 0.08% , 0.20% ) | 0.13% ( 0.08% , 0.20% ) | 111 | 14,598 | 0.76% ( 0.63% , 0.92% ) | 0.94% ( 0.76% , 1.16% ) |
|  | South West | 7 | 11,779 | 0.06% ( 0.02% , 0.12% ) | 0.05% ( 0.02% , 0.12% ) | 39 | 10,867 | 0.36% ( 0.26% , 0.49% ) | 0.43% ( 0.30% , 0.61% ) |

\* For categories other than age and region, we present weighted prevalence if the number of positives in a category is 10 or more.
\*\* Small number reporting 3 doses have been included in this group (<30 participants)

**Table 3b.**
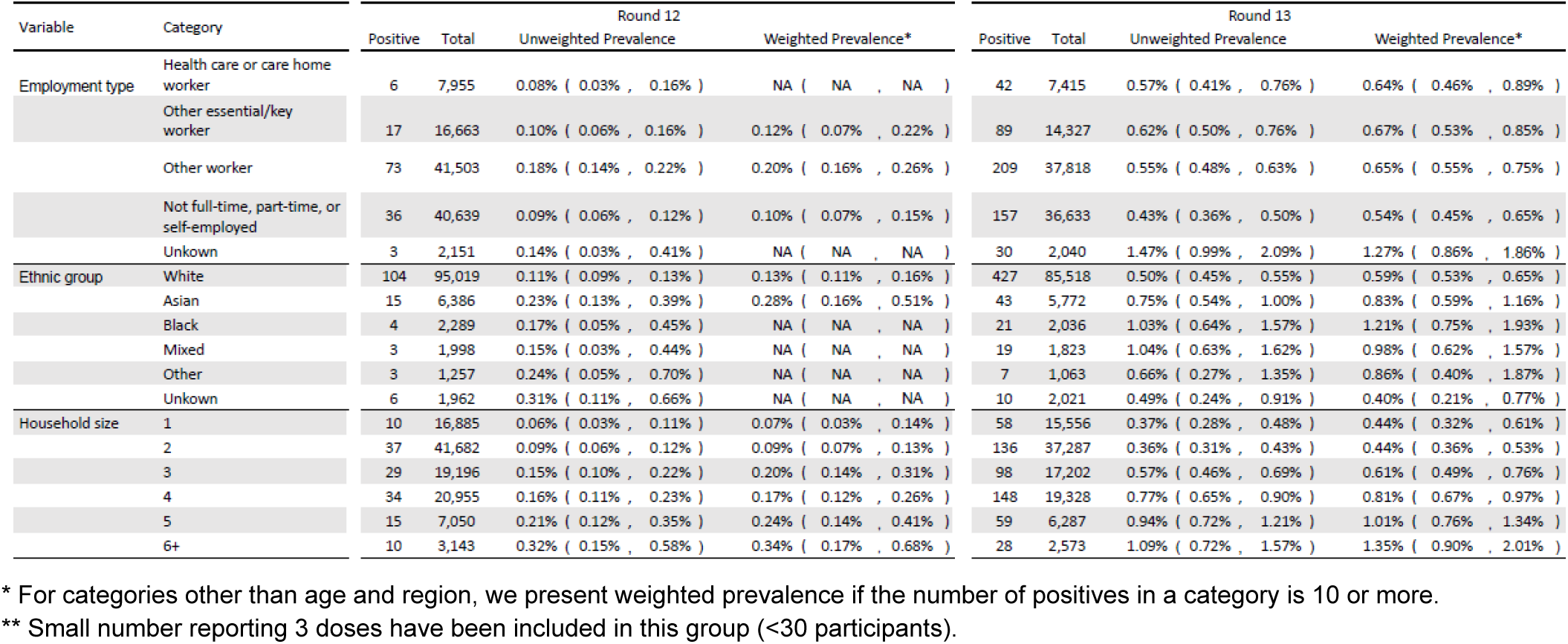
Unweighted and weighted prevalence of swab-positivity for employment type, ethnic group, and household size for round 12 and round 13.

**Table 3c.**
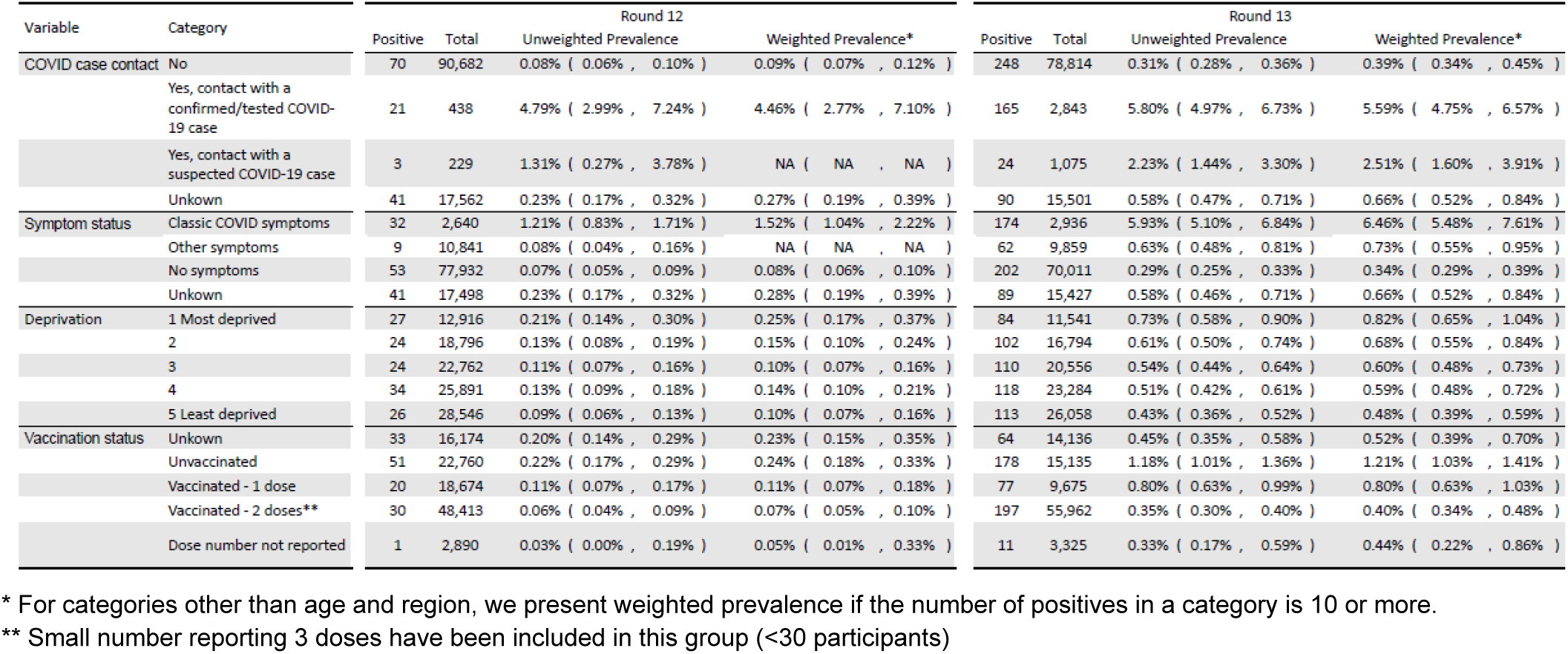
Unweighted and weighted prevalence of swab-positivity for COVID case contact status, symptom status, neighbourhood deprivation and vaccination status for round 12 and round 13.

**Figure 2.**
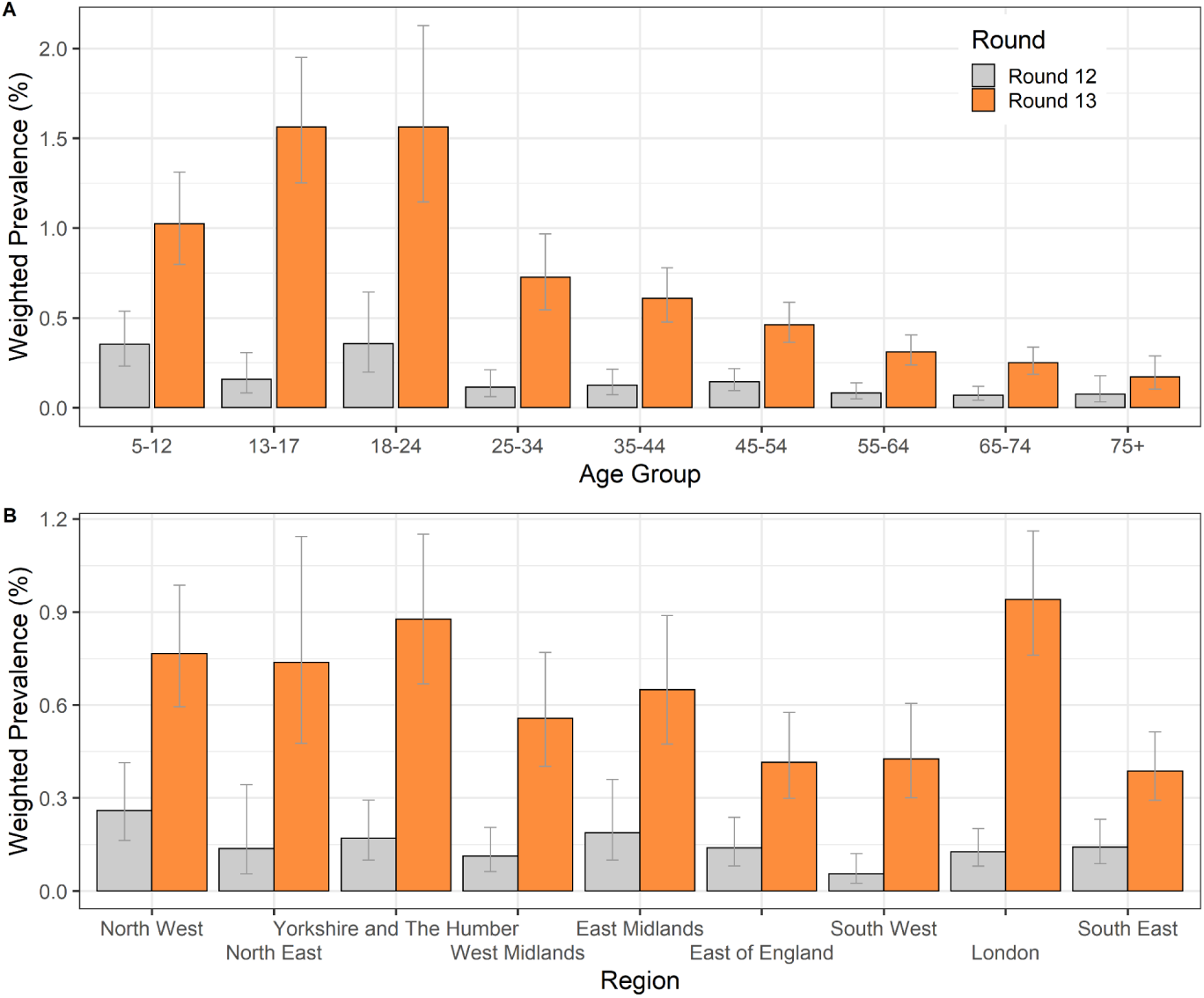
Weighted prevalence of swab-positivity by age group and by region for round 12 and round 13 complete. Bars show 95% confidence intervals. (A) Weighted prevalence of swab-positivity by age group. (B) Weighted prevalence of swab-positivity by region.

### Vaccine effectiveness

Participants who reported being vaccinated were at substantially reduced risk of testing positive compared with those who reported not being vaccinated. For round 13, prevalence of swab positivity among those unvaccinated was three-fold greater for all ages at 1.21% (1.03%, 1.41%) compared with 0.40% (0.34%, 0.48%) among those reporting two doses of vaccine (Table 3). The ratio of prevalence for unvaccinated to vaccinated individuals for round 12 was similar with a prevalence of 0.24% (0.18%, 0.33%) in those unvaccinated compared with 0.07% (0.05%, 0.10%) in those reporting two doses (Table 3).

However, these estimates conflate the effect of vaccination with other correlated variables such as age, which is strongly associated with likelihood of having been vaccinated and also acts as a proxy for differences in behaviour across the age groups. Specifically, in England, few children and young people under the age of 18 years have been vaccinated, while few over the age of 65 years remain unvaccinated (Table 4, Figure 1). We therefore restricted the analyses to those aged 18 to 64 years (n = 64,415 in round 12, n = 57,457 in round 13), which permitted direct contrast of infection rates between vaccinated and unvaccinated groups (Table 4a).

**Table 4.** Vaccination and swab-positivity status in rounds 12 and 13 of REACT 1 shown for all participants (5 years and over) and for the subset aged 18 to 64 years.

| Data set | Age group | Vaccine status | Round 12 |  |  | Round 13 |  |  |
| --- | --- | --- | --- | --- | --- | --- | --- | --- |
|  |  |  | Negative | Positive | Odds ratio | Negative | Positive | Odds ratio |
| Self-reported | All | Unvaccinated | 22,709 | 51 | Reference | 14957 | 178 | Reference |
|  |  | Vaccinated (1 dose) | 18,654 | 20 | 0.48 ( 0.28 , 0.80 ) | 9598 | 77 | 0.67 ( 0.52 , 0.88 ) |
|  |  | Vaccinated (2 or more doses) | 48,383 | 30 | 0.28 ( 0.18 , 0.43 ) | 55765 | 197 | 0.30 ( 0.24 , 0.36 ) |
|  |  | Vaccinated (unknown doses) | 2,889 | 1 | 0.15 ( 0.02 , 1.12 ) | 3314 | 11 | 0.28 ( 0.15 , 0.51 ) |
|  |  | Vaccine status not known | 16,141 | 33 | 0.91 ( 0.59 , 1.41 ) | 14072 | 64 | 0.38 ( 0.29 , 0.51 ) |
|  | 18-64 | Unvaccinated | 9,012 | 16 | Reference | 2574 | 28 | Reference |
|  |  | Vaccinated (1 dose) | 18,307 | 19 | 0.58 ( 0.30 , 1.14 ) | 9467 | 76 | 0.74 ( 0.48 , 1.14 ) |
|  |  | Vaccinated (2 or more doses) | 25,248 | 17 | 0.38 ( 0.19 , 0.75 ) | 34503 | 145 | 0.39 ( 0.26 , 0.58 ) |
|  |  | Vaccinated (unknown doses) | 1,173 | 0 | 0.00 ( 0.00 , NA ) | 1517 | 9 | 0.55 ( 0.26 , 1.16 ) |
|  |  | Vaccine status not known | 10,597 | 26 | 1.38 ( 0.74 , 2.58 ) | 9089 | 49 | 0.50 ( 0.31 , 0.79 ) |
| Linked | All | Unvaccinated | 19,115 | 52 | Reference | 11,357 | 153 | Reference |
|  |  | Vaccinated (1 dose) | 26,285 | 33 | 0.46 ( 0.30 , 0.71 ) | 11,885 | 93 | 0.58 ( 0.45 , 0.75 ) |
|  |  | Vaccinated (2 or more doses) | 50,721 | 34 | 0.25 ( 0.16 , 0.38 ) | 61,202 | 206 | 0.25 ( 0.20 , 0.31 ) |
|  | 18-64 | Unvaccinated | 8,099 | 21 | Reference | 1,553 | 25 | Reference |
|  |  | Vaccinated (1 dose) | 25,657 | 32 | 0.48 ( 0.28 , 0.83 ) | 11,652 | 92 | 0.49 ( 0.31 , 0.77 ) |
|  |  | Vaccinated (2 or more doses) | 23,511 | 18 | 0.30 ( 0.16 , 0.55 ) | 36,448 | 153 | 0.26 ( 0.17 , 0.40 ) |
|  | All | Unvaccinated | 19,115 | 52 | Reference | 11,357 | 153 | Reference |
|  |  | Vaccinated (< 14 days 2nd dose) | 31,826 | 35 | 0.40 ( 0.26 , 0.62 ) | 13,425 | 102 | 0.56 ( 0.44 , 0.73 ) |
|  |  | Vaccinated (>= 14 days 2nd dose) | 45,180 | 32 | 0.26 ( 0.17 , 0.40 ) | 59,662 | 197 | 0.25 ( 0.20 , 0.30 ) |
|  | 18-64 | Unvaccinated | 8,099 | 21 | Reference | 1,553 | 25 | Reference |
|  |  | Vaccinated (< 14 days 2nd dose) | 30,593 | 34 | 0.43 ( 0.25 , 0.74 ) | 13,170 | 101 | 0.48 ( 0.31 , 0.74 ) |
|  |  | Vaccinated (>= 14 days 2nd dose) | 18,575 | 16 | 0.33 ( 0.17 , 0.64 ) | 34,930 | 144 | 0.26 ( 0.17 , 0.39 ) |

At these ages, we compared swab-negatives with i) all swab-positives and ii) the subset of swab-positives who were symptomatic, that is reporting one or more classic COVID-19 symptoms in the month prior to testing (fever, loss or change of sense of smell or taste, new persistent cough). After adjusting for age, sex, region, ethnicity and index of multiple deprivation (IMD) [14], for all swab-positives, we estimated vaccine effectiveness (VE) in round 12 of 64% (11%, 85%) and 49% (22%, 67%) in round 13. For those with symptoms we estimated VE of 83% (19%, 97%) in round 12 and 59% (23%, 78%) in round 13.

In secondary analyses, for the 87% of participants aged 18 to 64 in round 13 who consented to data linkage (Methods), we estimated adjusted VE at 75% (35%, 90%) in round 12 and 62% (38%, 77%) in round 13. The apparent increase in VE for the linked participants reflected differences in odds of infection among the linked and unlinked groups (Table 5a), suggesting likely selection bias for consent to linkage. However, since the linked group had more reliable reported dates of vaccination, we examined the potential effect of a lag period of 14 days after the second vaccination and observed similar odds ratios for zero lag and 14 days lag following the second dose (Table 4).

**Table 5.**
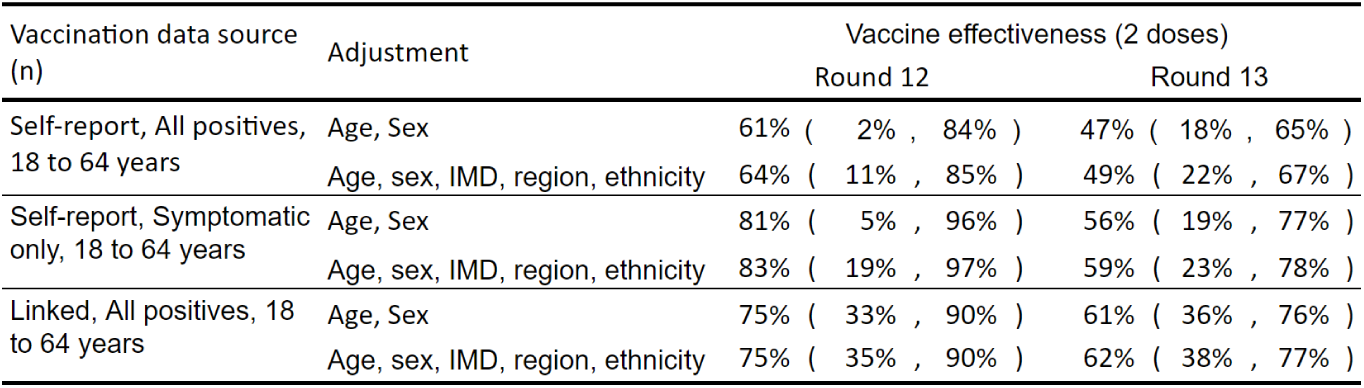
Unadjusted and adjusted estimates of vaccine effectiveness against infection for self-reported vaccine status and linked vaccine status for rounds 12 and 13 of REACT-1 for participants aged 18 to 64 years.

| Vaccination data source<br>(n) | Adjustment | Vaccine effectiveness (2 doses) |  |
| --- | --- | --- | --- |
|  |  | Round 12 | Round 13 |
| Self-report, All positives, 18 to 64 years | Age, Sex | 61% ( 2% , 84% ) | 47% ( 18% , 65% ) |
|  | Age, sex, IMD, region, ethnicity | 64% ( 11% , 85% ) | 49% ( 22% , 67% ) |
| Self-report, Symptomatic only, 18 to 64 years | Age, Sex | 81% ( 5% , 96% ) | 56% ( 19% , 77% ) |
|  | Age, sex, IMD, region, ethnicity | 83% ( 19% , 97% ) | 59% ( 23% , 78% ) |
| Linked, All positives, 18 to 64 years | Age, Sex | 75% ( 33% , 90% ) | 61% ( 36% , 76% ) |
|  | Age, sex, IMD, region, ethnicity | 75% ( 35% , 90% ) | 62% ( 38% , 77% ) |

**Table 5a.** Comparison self-reported vaccination status and swab-positivity for participants who did and did not consent to data linkage.

| Vaccination status | Linked |  |  | Unlinked |  |  |
| --- | --- | --- | --- | --- | --- | --- |
|  | Negative | Positive | Odds ratio | Negative | Positive | Odds ratio |
| Unvaccinated | 2174 | 20 | Reference | 400 | 8 | Reference |
| Vaccinated (1 dose) | 8048 | 67 | 90% ( 55% , 149% ) | 1419 | 9 | 32% ( 12% , 83% ) |
| Vaccinated (2 or more doses) | 30371 | 133 | 48% ( 30% , 76% ) | 4132 | 12 | 15% ( 6% , 36% ) |
| Vaccinated (unknown doses) | 1330 | 7 | 57% ( 24% , 136% ) | 187 | 2 | 53% ( 11% , 254% ) |
| Vaccine status not known | 7730 | 43 | 60% ( 35% , 103% ) | 1359 | 6 | 22% ( 8% , 64% ) |
| Total | 49653 | 270 |  | 7497 | 37 |  |

While vaccination was associated with lower prevalence of swab-positivity, there remained potential for large numbers of fully vaccinated people to become infected. During the period of round 12, we extrapolated from our data that 29% of infections in England occurred in double-vaccinated people, rising to 44% during the period of round 13. Also, although lower than for unvaccinated individuals, nearly one in 25 double-vaccinated individuals (3.84% [2.81%, 5.21%]) tested swab-positive if they reported contact with a known COVID-19 case (Table 6).

**Table 6.**
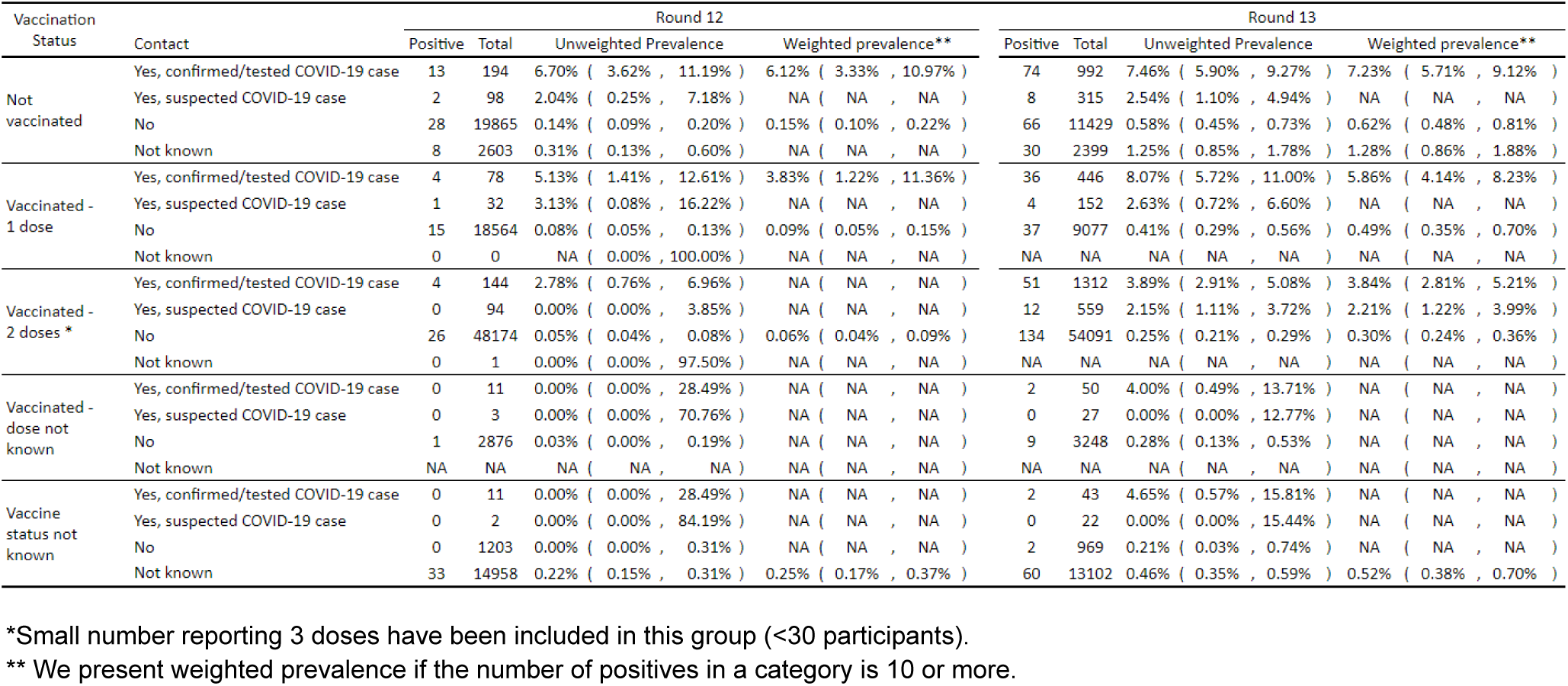
Unweighted and weighted prevalence of infection by vaccine and contact status for rounds 12 and 13 of REACT-1.

| Vaccination Status | Contact | Round 12 |  |  |  | Round 13 |  |  |  |
| --- | --- | --- | --- | --- | --- | --- | --- | --- | --- |
|  |  | Positive | Total | Unweighted Prevalence | Weighted prevalence** | Positive | Total | Unweighted Prevalence | Weighted prevalence** |
| Not vaccinated | Yes, confirmed/tested COVID-19 case | 13 | 194 | 6.70% ( 3.62% , 11.19% ) | 6.12% ( 3.33% , 10.97% ) | 74 | 992 | 7.46% ( 5.90% , 9.27% ) | 7.23% ( 5.71% , 9.12% ) |
|  | Yes, suspected COVID-19 case | 2 | 98 | 2.04% ( 0.25% , 7.18% ) | NA ( NA , NA ) | 8 | 315 | 2.54% ( 1.10% , 4.94% ) | NA ( NA , NA ) |
|  | No | 28 | 19865 | 0.14% ( 0.09% , 0.20% ) | 0.15% ( 0.10% , 0.22% ) | 66 | 11429 | 0.58% ( 0.45% , 0.73% ) | 0.62% ( 0.48% , 0.81% ) |
|  | Not known | 8 | 2603 | 0.31% ( 0.13% , 0.60% ) | NA ( NA , NA ) | 30 | 2399 | 1.25% ( 0.85% , 1.78% ) | 1.28% ( 0.86% , 1.88% ) |
| Vaccinated - 1 dose | Yes, confirmed/tested COVID-19 case | 4 | 78 | 5.13% ( 1.41% , 12.61% ) | 3.83% ( 1.22% , 11.36% ) | 36 | 446 | 8.07% ( 5.72% , 11.00% ) | 5.86% ( 4.14% , 8.23% ) |
|  | Yes, suspected COVID-19 case | 1 | 32 | 3.13% ( 0.08% , 16.22% ) | NA ( NA , NA ) | 4 | 152 | 2.63% ( 0.72% , 6.60% ) | NA ( NA , NA ) |
|  | No | 15 | 18564 | 0.08% ( 0.05% , 0.13% ) | 0.09% ( 0.05% , 0.15% ) | 37 | 9077 | 0.41% ( 0.29% , 0.56% ) | 0.49% ( 0.35% , 0.70% ) |
|  | Not known | 0 | 0 | NA ( 0.00% , 100.00% ) | NA ( NA , NA ) | NA | NA | NA ( NA , NA ) | NA ( NA , NA ) |
| Vaccinated - 2 doses * | Yes, confirmed/tested COVID-19 case | 4 | 144 | 2.78% ( 0.76% , 6.96% ) | NA ( NA , NA ) | 51 | 1312 | 3.89% ( 2.91% , 5.08% ) | 3.84% ( 2.81% , 5.21% ) |
|  | Yes, suspected COVID-19 case | 0 | 94 | 0.00% ( 0.00% , 3.85% ) | NA ( NA , NA ) | 12 | 559 | 2.15% ( 1.11% , 3.72% ) | 2.21% ( 1.22% , 3.99% ) |
|  | No | 26 | 48174 | 0.05% ( 0.04% , 0.08% ) | 0.06% ( 0.04% , 0.09% ) | 134 | 54091 | 0.25% ( 0.21% , 0.29% ) | 0.30% ( 0.24% , 0.36% ) |
|  | Not known | 0 | 1 | 0.00% ( 0.00% , 97.50% ) | NA ( NA , NA ) | NA | NA | NA ( NA , NA ) | NA ( NA , NA ) |
| Vaccinated - dose not known | Yes, confirmed/tested COVID-19 case | 0 | 11 | 0.00% ( 0.00% , 28.49% ) | NA ( NA , NA ) | 2 | 50 | 4.00% ( 0.49% , 13.71% ) | NA ( NA , NA ) |
|  | Yes, suspected COVID-19 case | 0 | 3 | 0.00% ( 0.00% , 70.76% ) | NA ( NA , NA ) | 0 | 27 | 0.00% ( 0.00% , 12.77% ) | NA ( NA , NA ) |
|  | No | 1 | 2876 | 0.03% ( 0.00% , 0.19% ) | NA ( NA , NA ) | 9 | 3248 | 0.28% ( 0.13% , 0.53% ) | NA ( NA , NA ) |
|  | Not known | NA | NA | NA ( NA , NA ) | NA ( NA , NA ) | NA | NA | NA ( NA , NA ) | NA ( NA , NA ) |
| Vaccine status not known | Yes, confirmed/tested COVID-19 case | 0 | 11 | 0.00% ( 0.00% , 28.49% ) | NA ( NA , NA ) | 2 | 43 | 4.65% ( 0.57% , 15.81% ) | NA ( NA , NA ) |
|  | Yes, suspected COVID-19 case | 0 | 2 | 0.00% ( 0.00% , 84.19% ) | NA ( NA , NA ) | 0 | 22 | 0.00% ( 0.00% , 15.44% ) | NA ( NA , NA ) |
|  | No | 0 | 1203 | 0.00% ( 0.00% , 0.31% ) | NA ( NA , NA ) | 2 | 969 | 0.21% ( 0.03% , 0.74% ) | NA ( NA , NA ) |
|  | Not known | 33 | 14958 | 0.22% ( 0.15% , 0.31% ) | 0.25% ( 0.17% , 0.37% ) | 60 | 13102 | 0.46% ( 0.35% , 0.59% ) | 0.52% ( 0.38% , 0.70% ) |

\*Small number reporting 3 doses have been included in this group (<30 participants).
\*\* We present weighted prevalence if the number of positives in a category is 10 or more.

### Cycle threshold values

We analysed Ct values associated with positive results among vaccinated and unvaccinated individuals as a measure of viral load and as a proxy for infectiousness. For all positives in round 13, at ages 18 to 64 years, median Ct value was higher for vaccinated participants at 27.6 (25.5, 29.7) compared with unvaccinated at 23.1 (20.3, 25.8) (positive defined as N gene Ct <37 or both N gene and E gene positive, Methods) (Table 7, Figure 3). The higher Ct values among vaccinated people indicate lower infectiousness, consistent with transmission studies conducted when the Alpha variant was dominant, in which vaccinated individuals were at substantially lower risk of passing on infection [15]. However, as a secondary analysis, we reduced the Ct threshold for positivity, representing strong positives with greater infectiousness. As the Ct threshold for positivity was reduced, the difference between medians for vaccinated and unvaccinated individuals became smaller. However, at the same time our estimate of vaccine effectiveness increased to 54% (29%, 71%) at a Ct threshold of 35, plateauing between 57% (32%, 72%) at a Ct threshold of 33 and 58% (33%, 73%) at a Ct threshold of 27 (Table 7, Figure 3).

**Table 7.** Median N-gene Ct values between individuals with different vaccine status, ages 18-64 years, round 13. Estimates have been calculated for all data available and subsets of data with lower N-gene Ct values.

| Data | Vaccine status | Negatives | Positives | Median N-gene Ct value* | P-value** |
| --- | --- | --- | --- | --- | --- |
| All data | Not vaccinated | 2574 | 28 | 23.1 ( 20.3 , 25.8 ) | ref |
|  | 1 vaccine doses | 9467 | 76 | 27.4 ( 24.8 , 30.0 ) | 0.04 |
|  | 2 vaccine doses | 34484 | 145 | 27.6 ( 25.5 , 29.7 ) | 0.01 |
|  | Vaccine doses not known | 1517 | 9 | 29.7 ( 25.6 , 33.7 ) | 0.01 |
| N-gene Ct <35 | Not vaccinated | 2574 | 27 | 22.9 ( 20.8 , 25.0 ) | ref |
|  | 1 vaccine doses | 9467 | 68 | 25.9 ( 23.6 , 28.2 ) | 0.09 |
|  | 2 vaccine doses | 34484 | 118 | 25.5 ( 23.9 , 27.1 ) | 0.09 |
|  | Vaccine doses not known | 1517 | 7 | 28.7 ( 26.5 , 31.0 ) | 0.05 |
| N-gene Ct <33 | Not vaccinated | 2574 | 26 | 22.9 ( 20.4 , 25.5 ) | ref |
|  | 1 vaccine doses | 9467 | 62 | 25.2 ( 22.6 , 27.8 ) | 0.15 |
|  | 2 vaccine doses | 34484 | 99 | 24.3 ( 22.5 , 26.1 ) | 0.41 |
|  | Vaccine doses not known | 1517 | 6 | 27.7 ( 24.3 , 31.2 ) | 0.10 |

\* 95% Confidence intervals in the median were estimated using quantile regression
\*\*P-values were calculated using the two-sided Wilcoxon two-sample test (also known as Mann Whitney-U) and are relative to the distribution of Ct values in individuals who are not vaccinated

**Figure 3.**
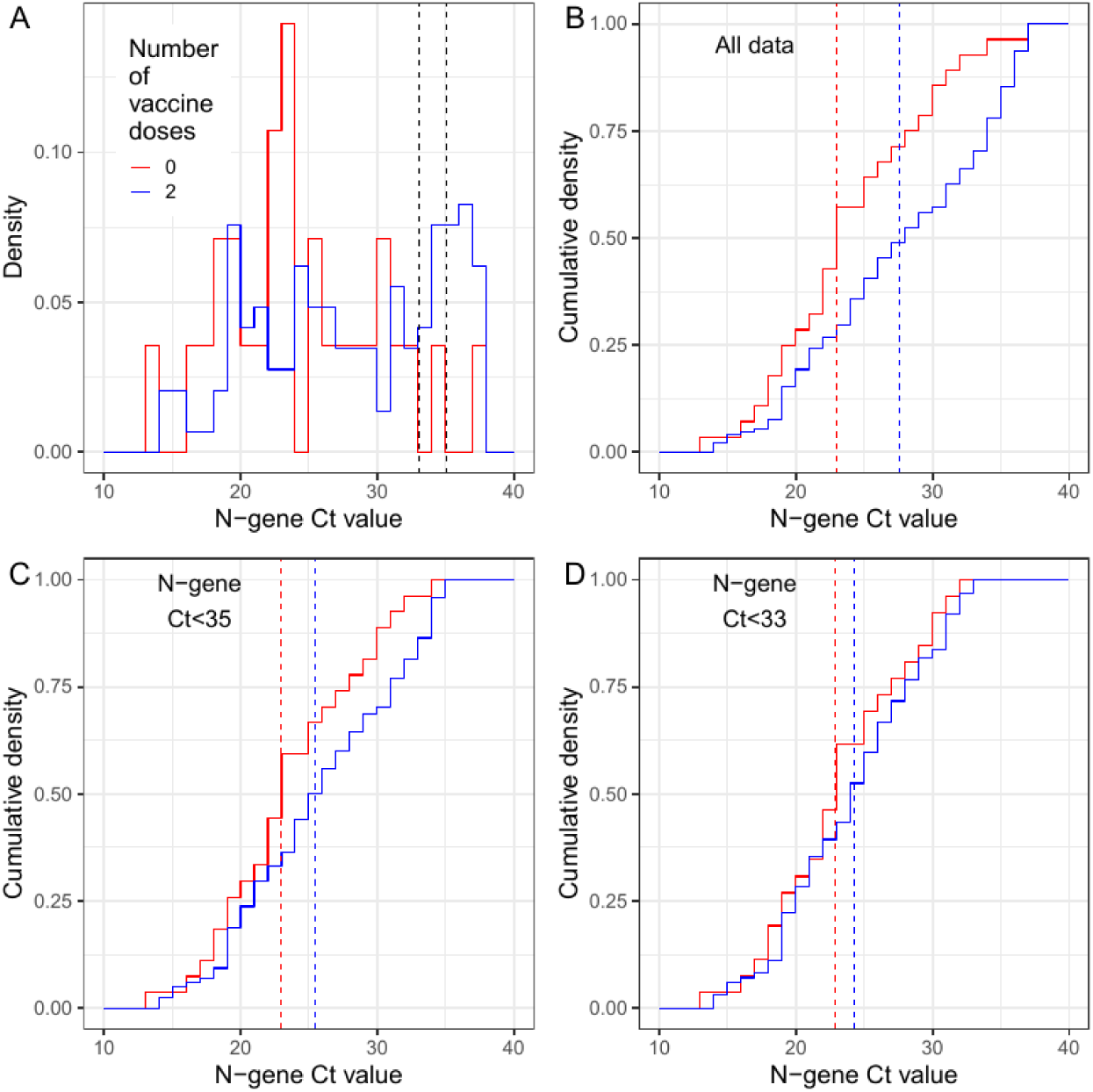
Distribution of N-gene Ct values, by vaccine status, for positive samples obtained from individuals aged 18-64 years inclusive. (A) Distribution of all N-gene Ct values for those who are unvaccinated (red) and those who reported receiving two doses of a vaccine (blue). Also shown are two black dotted lines at N-gene Ct equals 35 and N-gene Ct equals 33; these show the threshold values for a sample to be classed as positive used in sensitivity analyses. (B) Cumulative density of N-gene Ct values using all available data for unvaccinated individuals (red) and individuals who have had two doses of a vaccine (blue). (B) Cumulative density of N-gene Ct values using all data in which N-gene Ct is less than 35 for unvaccinated individuals (red) and individuals who have had two doses of a vaccine (blue). (D) Cumulative density of N-gene Ct values using all data in which N-gene Ct is less than 33 for unvaccinated individuals (red) and individuals who have had two doses of a vaccine (blue). Red and blue vertical dashed lines show the median value for each distribution.

### Link between time series of infections, hospital admissions and deaths

We next investigated how swab-positivity measured in REACT-1 related to daily hospital admissions and deaths in public data [6], finding a best fitting lag between swab-positivity and hospitalisations of 20 days and between swab-positivity and deaths of 26 days (Figure 5). At these lags, from early February 2021, there was a clear divergence between swab-positivity and deaths, with a smaller divergence between swab-positivity and hospitalisations, coinciding with the roll-out of England’s mass vaccination campaign. However, as the Delta variant became dominant in mid-April 2021, the associations between infections and hospitalisations and deaths began to re-converge, both for people below and above 65 years (Figure 6).

### Geographical variation

At the regional level, estimates of R were consistent with the overall trend within round 13. Prevalence in round 13 was highest in London at 0.94% (0.76%, 1.16%) up from 0.13% (0.08%, 0.20%) in round 12 (Table 3). There was a suggestion of a possible slowing of the rise in London in the most recent data, although with wide confidence intervals (Table 8).

**Table 8.** Estimates of regional growth rates, doubling times and reproduction numbers for round 13, and round 12 to round 13.

| Round | Region | Growth rate | R | Probability<br>R>1 | Halving (-) / Doubling (+)<br>time |
| --- | --- | --- | --- | --- | --- |
| 12 and 13 | East Midlands | 0.038 ( 0.024 , 0.055 ) | 1.26 ( 1.16 , 1.38 ) | >0.99 | 18.0 ( 29.5 , 12.6 ) |
|  | West Midlands | 0.048 ( 0.031 , 0.066 ) | 1.33 ( 1.21 , 1.47 ) | >0.99 | 14.5 ( 22.3 , 10.5 ) |
|  | East of England | 0.032 ( 0.017 , 0.048 ) | 1.21 ( 1.11 , 1.33 ) | >0.99 | 21.9 ( 41.9 , 14.4 ) |
|  | London | 0.052 ( 0.040 , 0.064 ) | 1.36 ( 1.28 , 1.46 ) | >0.99 | 13.4 ( 17.2 , 10.8 ) |
|  | North West | 0.031 ( 0.021 , 0.043 ) | 1.21 ( 1.14 , 1.29 ) | >0.99 | 22.1 ( 33.3 , 16.2 ) |
|  | North East | 0.050 ( 0.029 , 0.076 ) | 1.35 ( 1.19 , 1.55 ) | >0.99 | 13.8 ( 24.0 , 9.1 ) |
|  | South East | 0.029 ( 0.017 , 0.042 ) | 1.19 ( 1.11 , 1.29 ) | >0.99 | 24.3 ( 42.0 , 16.6 ) |
|  | South West | 0.058 ( 0.037 , 0.083 ) | 1.41 ( 1.25 , 1.61 ) | >0.99 | 11.9 ( 18.8 , 8.3 ) |
|  | Yorkshire ** | 0.046 ( 0.033 , 0.061 ) | 1.32 ( 1.22 , 1.43 ) | >0.99 | 14.9 ( 21.1 , 11.3 ) |
| 13 | East Midlands | 0.054 ( -0.009 , 0.117 ) | 1.38 ( 0.94 , 1.91 ) | 0.95 | 12.8 ( * , 5.9 ) |
|  | West Midlands | 0.063 ( 0.001 , 0.127 ) | 1.45 ( 1.01 , 2.00 ) | 0.98 | 10.9 ( * , 5.5 ) |
|  | East of England | 0.014 ( -0.055 , 0.081 ) | 1.09 ( 0.69 , 1.59 ) | 0.66 | * ( -12.7 , 8.6 ) |
|  | London | -0.017 ( -0.056 , 0.022 ) | 0.90 ( 0.68 , 1.15 ) | 0.20 | -41.5 ( -12.3 , 31.6 ) |
|  | North West | 0.045 ( -0.003 , 0.092 ) | 1.31 ( 0.98 , 1.69 ) | 0.97 | 15.4 ( * , 7.5 ) |
|  | North East | 0.031 ( -0.051 , 0.112 ) | 1.21 ( 0.71 , 1.86 ) | 0.77 | 22.1 ( -13.6 , 6.2 ) |
|  | South East | 0.017 ( -0.042 , 0.074 ) | 1.11 ( 0.75 , 1.54 ) | 0.71 | 41.8 ( -16.5 , 9.3 ) |
|  | South West | 0.049 ( -0.020 , 0.120 ) | 1.34 ( 0.88 , 1.93 ) | 0.92 | 14.3 ( -34.3 , 5.8 ) |
|  | Yorkshire ** | 0.042 ( -0.009 , 0.093 ) | 1.29 ( 0.95 , 1.69 ) | 0.95 | 16.4 ( -79.4 , 7.5 ) |

\* Doubling/Halving time had an estimated magnitude greater than 50 days and so represented approximately constant prevalence
\*\* Yorkshire and The Humber

At the sub-regional level, there was a suggestion of prevalence of infection decreasing in some areas and increasing in others (Figure 4). For example, in the North West of England, high prevalence in a large urban area covering Greater Manchester and Lancashire during the first half of round 13 was less evident in the second half, whereas prevalence increased between the first and second halves in nearby south Yorkshire, part of the Yorkshire and The Humber region. These data are indicative of rapidly changing local spread of the virus within the context of the national exponential rise in infections.

**Figure 4.**
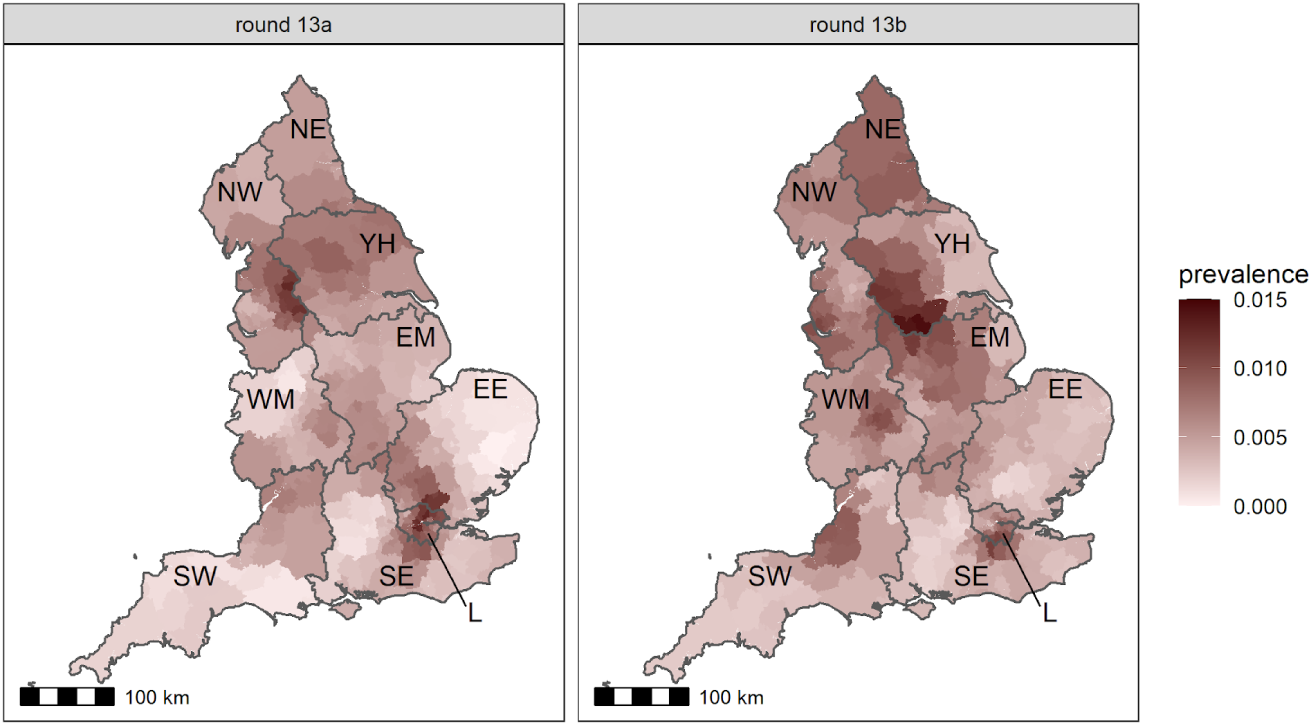
Neighbourhood smoothed average prevalence by lower tier local area for (A) round 13a and (B) round 13b. Neighbourhood prevalence calculated from nearest neighbours (the median number of neighbours within 30 km in the study). Average neighbourhood prevalence displayed for individual lower-tier local authorities. Regions: NE = North East, NW = North West, YH = Yorkshire and The Humber, EM = East Midlands, WM = West Midlands, EE = East of England, L = London, SE = South East, SW = South West.

**Figure 5.**
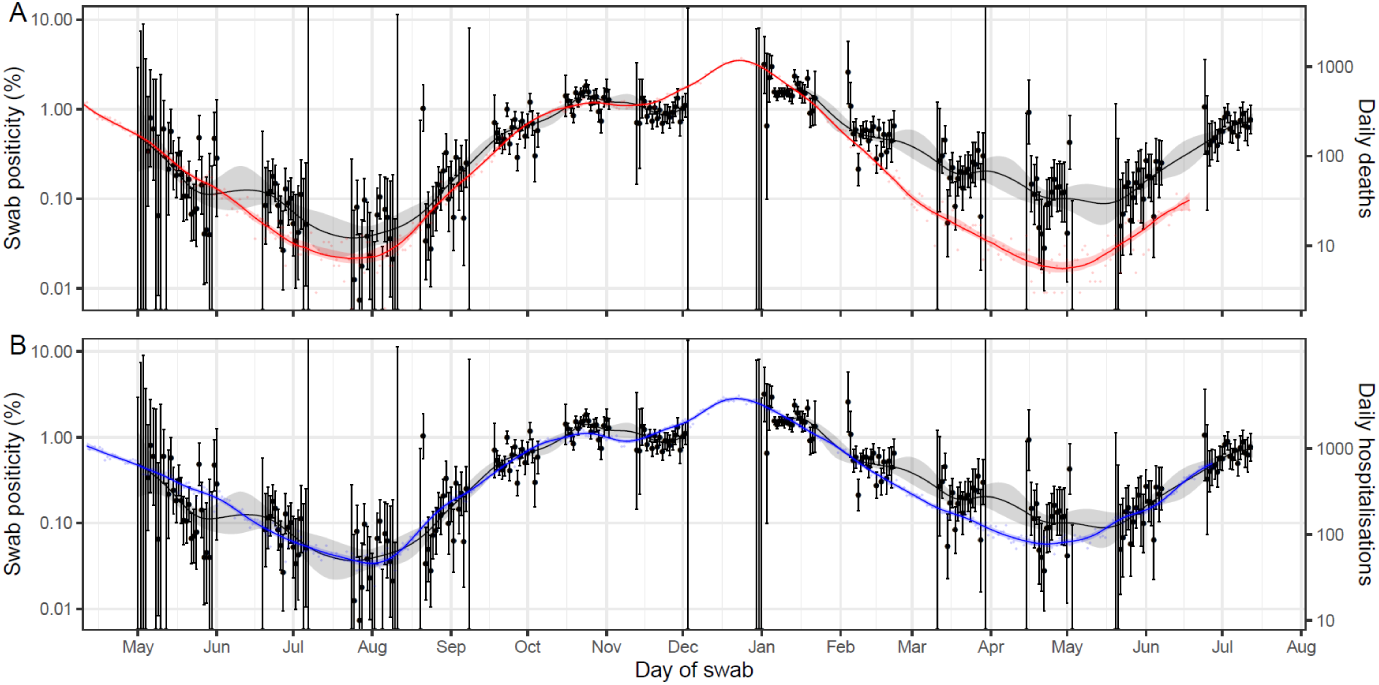
A comparison of daily deaths and hospitalisations to swab positivity as measured by REACT-1. Daily swab positivity for all 13 rounds of the REACT-1 study (black points with 95% confidence intervals, left hand y-axis) with P-spline estimates for swab positivity (solid black line, shaded area is 95% confidence interval). (A) Daily deaths in England (red points, right hand y-axis) and P-spline model estimates for expected daily deaths in England (solid red line, shaded area is 95% confidence interval, right hand y-axis). Daily deaths have been shifted by 26 (26, 26) days backwards in time along the x-axis. The two y-axes have been scaled using the best-fit population adjusted scaling parameter 0.059 (0.058, 0.061). (B) Daily hospitalisations in England (blue points, right hand y-axis) and P-spline model estimates for expected daily hospitalisations in England (solid blue line, shaded area is 95% confidence interval, right hand y-axis). Daily hospitalisations have been shifted by 20 (19, 20) days backwards in time along the x-axis. The two y-axes have been scaled using the best-fit population adjusted scaling parameter 0.241 (0.236, 0.246).

**Figure 6.**
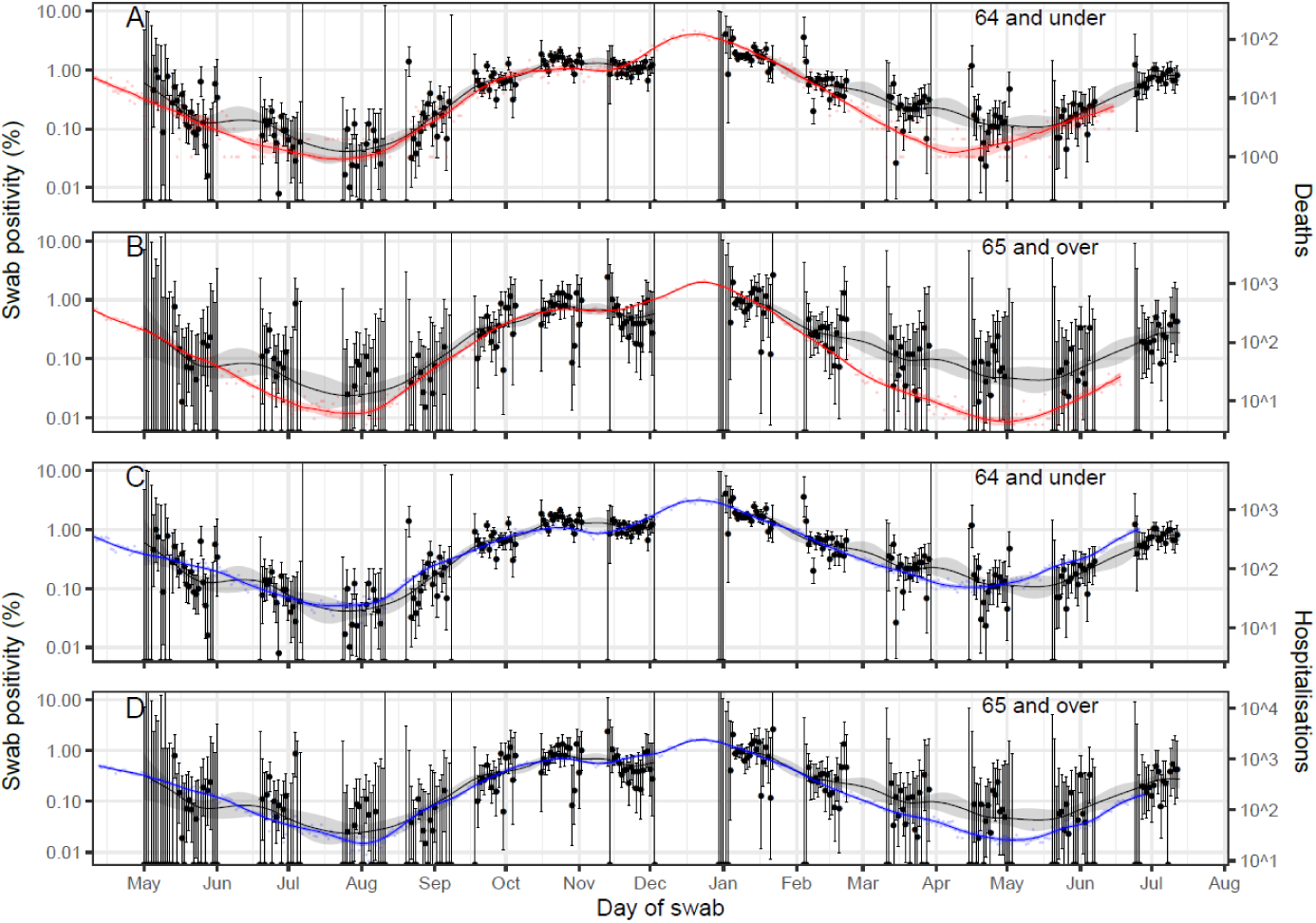
A comparison of daily deaths and hospitalisations to swab positivity as measured by REACT-1, by age group. Daily swab positivity for all 12 rounds of the REACT-1 study (black points with 95% confidence intervals, left hand y-axis) with P-spline estimates for swab positivity (solid black line, shaded area is 95% confidence interval) for (A, C) those aged under 64 years and (B,D) those aged 65 years and over. **(A)** Daily deaths for those aged 64 and under in England (red points, right hand y-axis) and corresponding P-spline model estimates for the expected number of deaths (solid red line, shaded area is 95% confidence interval, right hand y-axis). Daily deaths have been shifted by 29 (29, 29) days backwards in time along the x-axis. The two y-axes have been scaled using the best-fit population adjusted scaling parameter 0.0065 (0.0064, 0.0067). **(B)** Daily deaths for those aged 65 and over in England (red points, right hand y-axis) and corresponding P-spline model estimates for the expected number of deaths (solid red line, shaded area is 95% confidence interval, right hand y-axis). Daily deaths have been shifted by 26 (25, 28) days backwards in time along the x-axis. The two y-axes have been scaled using the best-fit scaling parameter 0.51 (0.48, 0.54). **(C)** Daily hospitalisations for those aged 64 and under in England (blue points, right hand y-axis) and corresponding P-spline model estimates for the expected number of hospitalisations (solid blue line, shaded area is 95% confidence interval, right hand y-axis). Daily hospitalisations have been shifted by 21 (20, 21) days backwards in time along the x-axis. The two y-axes have been scaled using the best-fit scaling parameter 0.101 (0.098, 0.103). **(D)** Daily hospitalisations for those aged 65 and over in England (blue points, right hand y-axis) and corresponding P-spline model estimates for the expected number of hospitalisations (solid blue line, shaded area is 95% confidence interval, right hand y-axis). Daily hospitalisations have been shifted by 19 (17, 20) days backwards in time along the x-axis. The two y-axes have been scaled using the best-fit scaling parameter 1.41 (1.33, 1.50).

### Sex, ethnicity, household size and neighbourhood deprivation

Ethnicity, household size and area levels of deprivation jointly contributed to the risk of higher prevalence of swab-positivity. Unadjusted prevalence (Table 3) showed: highest prevalence in people of Black ethnicity at 1.21% (0.75%, 1.93%) compared with 0.59% (0.53%, 0,65%) in people of white ethnicity; highest prevalence in those in the largest households of 6 or more people at 1.35% (0.90%, 2.01%) compared with 0.44% (0.32%, 0.61%) and 0.44% (0.36%, 0.53%) in single and two person households respectively; and highest prevalence in participants living in the most deprived neighbourhoods at 0.82% (0.65%, 1.04%) compared with the least deprived at 0.48% (0.39%, 0.59%). In models including each of the above variables, similar patterns were observed in the odds of testing positive, although odds were reduced when all three of the above variables were considered jointly (Table 9). Also, in mutually adjusted analyses, women had lower odds of infection than men at 0.80 (0.67, 0.96) in round 13, although not in round 12 at 1.34 (0.93, 1.92) (Table 3, Table 9); this difference is possibly related to increased social mixing associated with England’s progression in the Euro 2020 football competition during June and July 2021, as was seen earlier in Scottish data, reflecting their earlier exit from the competition [16].

**Table 9.** Multivariable logistic regression for rounds 12 and round 13.

| Variable | Category | Round 12** |  | Round 13 ** |  |
| --- | --- | --- | --- | --- | --- |
|  |  | Adjusted for age and sex | Mutually adjusted* | Adjusted for age and sex | Mutually adjusted* |
| Sex | Male | Ref | Ref | Ref | Ref |
|  | Female | 1.13 [0.80,1.59] | 1.34 [0.93,1.92] | 0.82 [0.69,0.97] | 0.80 [0.67,0.96] |
| Age Group | 5-12 | 2.73 [1.46,5.10] | 2.61 [1.32,5.17] | 1.60 [1.15,2.23] | 1.71 [1.22,2.41] |
|  | 13-17 | 1.34 [0.61,2.96] | 1.87 [0.76,4.63] | 2.58 [1.89,3.53] | 2.82 [1.90,4.18] |
|  | 18-24 | 2.33 [1.10,4.92] | 2.57 [1.18,5.64] | 2.50 [1.73,3.62] | 2.61 [1.78,3.83] |
|  | 25-34 | 0.95 [0.44,2.06] | 1.12 [0.50,2.47] | 1.12 [0.79,1.60] | 1.14 [0.79,1.63] |
|  | 35-44 | Ref | Ref | Ref | Ref |
|  | 45-54 | 1.03 [0.55,1.96] | 1.22 [0.62,2.39] | 0.76 [0.55,1.05] | 0.76 [0.54,1.06] |
|  | 55-64 | 0.60 [0.30,1.20] | 0.82 [0.39,1.72] | 0.50 [0.36,0.71] | 0.54 [0.38,0.78] |
|  | 65+ | 0.54 [0.28,1.04] | 0.80 [0.36,1.75] | 0.36 [0.26,0.50] | 0.40 [0.27,0.59] |
| Region | North East | 1.12 [0.45,2.78] | 1.14 [0.45,2.86] | 1.80 [1.14,2.84] | 1.81 [1.14,2.88] |
|  | North West | 1.54 [0.85,2.81] | 1.40 [0.75,2.63] | 2.00 [1.43,2.79] | 1.73 [1.21,2.47] |
|  | Yorkshire and The Humber | 1.37 [0.72,2.63] | 1.39 [0.71,2.70] | 2.12 [1.50,2.99] | 2.05 [1.43,2.92] |
|  | East Midlands | 1.18 [0.58,2.39] | 1.20 [0.58,2.47] | 1.63 [1.11,2.39] | 1.62 [1.10,2.39] |
|  | West Midlands | 0.99 [0.48,2.01] | 0.77 [0.36,1.66] | 1.42 [0.97,2.07] | 1.36 [0.92,2.00] |
|  | East of England | 1.24 [0.65,2.34] | 1.06 [0.54,2.11] | 1.05 [0.71,1.56] | 1.03 [0.69,1.55] |
|  | London | 1.07 [0.59,1.95] | 0.84 [0.44,1.60] | 1.92 [1.41,2.64] | 1.69 [1.20,2.36] |
|  | South East | Ref | Ref | Ref | Ref |
|  | South West | 0.55 [0.23,1.30] | 0.58 [0.25,1.38] | 1.05 [0.70,1.57] | 0.97 [0.64,1.49] |
| Key Worker Status | HCW/CHW | 0.37 [0.16,0.85] | 0.37 [0.16,0.87] | 1.00 [0.71,1.40] | 0.99 [0.70,1.39] |
|  | Key worker (other) | 0.52 [0.30,0.88] | 0.55 [0.32,0.94] | 1.07 [0.83,1.38] | 1.08 [0.83,1.39] |
|  | Other worker | Ref | Ref | Ref | Ref |
|  | Not FT, PT, SE | 0.58 [0.36,0.92] | 0.60 [0.38,0.95] | 0.88 [0.69,1.12] | 0.87 [0.68,1.11] |
| Ethnicity | White | Ref | Ref | Ref | Ref |
|  | Asian | 1.69 [0.97,2.93] | 1.52 [0.84,2.76] | 1.08 [0.78,1.48] | 1.01 [0.72,1.41] |
|  | Black | 1.27 [0.47,3.47] | 1.26 [0.45,3.55] | 1.57 [1.01,2.45] | 1.48 [0.93,2.35] |
|  | Mixed | 0.91 [0.29,2.91] | 0.95 [0.30,3.05] | 1.27 [0.80,2.03] | 1.00 [0.58,1.73] |
|  | Other | 1.85 [0.59,5.87] | 1.82 [0.57,5.87] | 1.09 [0.51,2.30] | 0.87 [0.39,1.98] |
| Household Size | 1-2 People | Ref | Ref | Ref | Ref |
|  | 3-5 People | 1.28 [0.81,2.01] | 1.19 [0.75,1.89] | 0.94 [0.75,1.17] | 0.95 [0.76,1.20] |
|  | 6+ People | 2.07 [0.97,4.41] | 1.74 [0.78,3.88] | 1.19 [0.78,1.83] | 1.14 [0.72,1.80] |
| Deprivation Index Quintile | 1 - Most Deprived | 2.06 [1.20,3.54] | 1.94 [1.08,3.51] | 1.48 [1.12,1.97] | 1.29 [0.95,1.75] |
|  | 2 | 1.32 [0.76,2.31] | 1.39 [0.78,2.51] | 1.31 [1.00,1.71] | 1.15 [0.86,1.53] |
|  | 3 | 1.15 [0.66,2.00] | 1.11 [0.62,2.01] | 1.20 [0.92,1.56] | 1.12 [0.85,1.48] |
|  | 4 | 1.45 [0.87,2.42] | 1.55 [0.91,2.63] | 1.17 [0.90,1.52] | 1.13 [0.86,1.47] |
|  | 5 - Least Deprived | Ref | Ref | Ref | Ref |

\* Odds ratios mutually adjusted for all variables shown.
\*\*The sample size for variables in round 12 is 104,844 and in round 13 is 94,223

## Discussion

We report a rapidly rising prevalence of infection in England during 20 May to 12 July 2021 associated with the replacement of Alpha by Delta variant, in a highly vaccinated population. Our estimate of vaccine effectiveness against all SARS-CoV-2 infections for two doses of vaccine was 49% in the most recent data, increasing to 58% when we defined effectiveness only for strong positives. These estimates are lower than some others [15,17,18], but consistent with more recent data from Israel [19]. Our estimates were higher when we restricted our analyses to people reporting symptoms of COVID-19 in the previous month. These higher estimates were still lower than those reported using a test-negative design for routine testing of symptomatic people presenting for RT-PCR in England [17]. However, our data are based on a random sample of the population and include asymptomatic people, as well as symptomatic individuals who may not present for routine testing, and may therefore give a less biased representation of transmission risk. Also, our estimated effectiveness was lower than that from a longitudinal household survey which included asymptomatic individuals but which was conducted prior to the emergence of Delta [15].

We show that the third wave of infections in England was being driven primarily by the Delta variant in younger, unvaccinated people. This focus of infection offers considerable scope for interventions to reduce transmission among younger people, with knock-on benefits across the entire population. Also, given the rapid rise of the Delta variant in Europe, the USA, South Asia and elsewhere, and its estimated increased transmissibility, patterns observed in England indicate what may happen elsewhere. In our data, the highest prevalence of infection was among 12 to 24 year olds, raising the prospect that vaccinating more of this group by extending the UK programme to those aged 12 to 17 years could substantially reduce transmission potential in the autumn when levels of social mixing increase [20]. Also, development of vaccines against Delta may be warranted in the light of evidence of antigenic change measured by neutralization [21] and the relationship between neutralization titre and protection from mild disease [22].

Estimates of effectiveness against serious outcomes of greater than 90% have been reported for those who have received two doses of either BNT162b2 [23] or ChAdOx1-S [24] vaccines. This is in keeping with our observation of a weakening of the association between infections and hospitalisations and deaths from mid-February to early April 2021 when Alpha variant was dominant. However, in our more recent data (since mid-April 2021), infections and hospitalisations began to re-converge, potentially reflecting the increased prevalence and severity of Delta compared with Alpha [25], a changing age mix of severe cases, and possible waning of protection [19,26].

Our study has limitations. Our primary estimate of effectiveness was based on self-reported vaccine status, because we could only obtain linked vaccination data for the subset of participants who gave consent and that group appeared to have different patterns of swab-positivity across the vaccinated and unvaccinated groups. Over the course of the study since round 1 in May 2020, towards the end of the first lockdown in England, we have observed a gradual reduction in response rates, from 30.5% in round 1 to 11.7% in round 13. These rates are conservative estimates since they are based on numbers of swabs with a valid RT-PCR result compared to the total number of letters of invitation sent out, some of which may have been returned, sent to the wrong address or left unopened by the recipient. Nonetheless, the drop in response rates means that our sample may be becoming less representative, particularly in some groups such as young people (18 to 24 years) and those living in the most deprived areas where response rates have fallen to 4.2% and 5.1% respectively. It should be noted, however, that these response rates have been achieved without use of financial or other incentives.

Our method of sampling was designed initially to achieve sufficient numbers in each lower-tier local authority (LTLA) in England so that we could analyse sub-regional trends and also, by weighting the sample, provide estimates of prevalence that were representative of the population of England. While previously we had aimed to achieve approximately equal numbers of people in our sample by LTLA, in rounds 12 and 13 we switched to sampling in proportion to population in order to capture greater resolution in inner city areas, which were relatively under-represented in our previous sampling regimen. In either case, as we re-weight the sample according to the national population profile, weighted prevalence should be comparable across rounds, albeit with lower precision in later rounds because of the lower response rates.

In conclusion, we have shown rapid exponential growth of SARS-CoV2 prevalence during the third wave in England at a time when Delta variant became dominant. The rapid roll-out of the vaccination programme in England has so far limited the number of infections and serious cases relative to the unvaccinated population. Level or declining prevalence may be observed during the summer in the northern hemisphere, reflecting school vacations, greater time spent outdoors and reduced social interactions. But without additional interventions, increased mixing during the autumn in the presence of Delta variant may lead to renewed growth, even at high levels of vaccination. Continued surveillance to monitor the spread of the epidemic is therefore required.

## Materials and Methods

The REACT study methods have been described elsewhere [9]. Briefly, in REACT-1, non-overlapping random cross-sectional samples of the population in England at ages 5 years and above were sent invitation letters through the post. Each round of data collection took place approximately monthly over a period of two to three weeks (except December 2020 when no survey was undertaken) (Table 1). At each round, we invited named individuals obtained from NHS Digital based on the National Health Service (NHS) list of patients registered with a general practitioner in England, covering almost the entire population. We included all 317 lower-tier local authorities (LTLAs) in England, and by combining the Isles of Scilly with Cornwall and the City of London with Westminster, we report results across 315 LTLAs overall.

For round 1 to round 11 we aimed to obtain approximately equal numbers of participants in each LTLA to be powered to provide local estimates of prevalence. From round 12 onwards, we adjusted the sampling procedure to select the sample randomly in proportion to population at LTLA level thus obtaining more samples in higher population density LTLAs in inner urban areas. However, we ensured that data were comparable across rounds as we re-weighted the data at each round to be representative of England as a whole (see below).

For those registering to participate, we obtained age, sex, address and residential postcode from the NHS register and collected additional information on demographics, health and lifestyle via online or telephone questionnaire. This included information on ethnicity, smoking, household size, key worker status, contact with a known or suspected COVID-19 case, and whether, at time of survey, participants had experienced one or more of 29 symptoms in the past week or past month (participants not reporting symptoms may have developed symptoms later but these were not captured). Participants were also asked for consent to longer-term follow-up through linkage to their NHS records including data from the national immunization programme. The questionnaires are available on the study website [27].

Response rates have varied by age and over time and place, and are available for each round (“For Researchers: REACT-1 Study Materials” [27]). Overall response rate was defined as the percentage of invitees from whom we received a valid swab result; this was 20.4% across all rounds, and 13.4% and 11.7% for rounds 12 and 13 respectively. In round 13, response rate varied by age from 4.2% at ages 18 to 24 years to 24% at ages 65 to 74 years and by IMD decile from 5.1% in the most deprived areas to 20.8% in the least deprived.

Participants were requested to provide a self-administered throat and nose swab (obtained by parent or guardian for children aged 5 to 12 years) following written and video instructions. Swabs were placed into a dry tube (no solution or preservative), refrigerated at home, picked up by courier and then sent chilled to a single commercial laboratory for testing for SARS-CoV-2 by RT-PCR.

### Ct threshold and laboratory calibration experiments

We tested two gene targets (E gene and N gene) with cycle threshold (Ct) values used as a proxy for intensity of viral load. The RT-PCR test was considered positive if both gene targets were detected or if N gene was detected with Ct value less than 37. The Ct threshold used to determine positivity was set following three separate calibration experiments. First,10 RNA extraction plates were sent from the commercial laboratory for blinded re-analysis in two laboratories accredited by the UK Accreditation Service (UKAS). We found concordant results for 919 negative samples and all 40 controls. We detected viral RNA in 11 of the 19 samples with a Ct value reported positive by the commercial laboratory (N gene Ct value ranging from 16.5 to 40.7); in 10 of these 11 samples, N gene Ct value was < 37. Second, in a serial dilution experiment of synthetic SARS-CoV-2 RNA the commercial laboratory detected 2.5 copies at Ct 38; also whilst following serial dilution of known positive samples with low viral load, the commercial laboratory identified an N gene signal at Ct > 37 in most instances. Third, a Public Health England (PHE) reference laboratory re-analysed a further 40 unblinded positive samples (on 19 × 96 well plates) with N gene Ct values > 35 (range 35.7 to 46.8) and without a signal for E gene, detecting SARS CoV-2 RNA in 15/40 (38%) samples (2/4 with N gene Ct value < 37). The results of all three calibration experiments were then consolidated to set the positivity criteria noted above, which have been used throughout each round of REACT-1.

### Prevalence estimates and weighting

We obtained unweighted (crude) prevalence estimates for different sociodemographic and occupational groups by dividing counts of swab-positivity (based on RT-PCR) by the number of swabs returned in that group. We then applied rim weighting [28] to provide prevalence weighted to be representative of the population of England as a whole, by: age, sex, deciles of the IMD, LTLA counts and ethnic group. We obtained the age by sex and LTLA counts from the Office for National Statistics mid-year population estimates [29], counts by ethnic group from the Labour Force Survey, and calculated the IMD decile points from linkage of postcode to area-level IMD using the original sampling frame obtained from NHS Digital. Because of the different sources of population estimates, the rim weighting was based on proportions rather than population totals. We grouped age into seven categories: 18 to 24; 25 to 34; 35 to 44; 45 to 54; 55 to 64; 65 to 74; 75 years or above, giving 14 age-sex categories. Self-reported ethnicity was grouped into nine categories: white; mixed / multiple ethnic groups; Indian; Pakistani; Bangladeshi; Chinese; any other Asian background; Black African / Caribbean / other; and any other ethnic group or missing.

For the rim weighting, initially (first stage) the sample was weighted to LTLA counts and age by sex groups only, adjusting the age and sex groups to ensure that the final weighted estimates were as close as possible to the population profile. Then, using the first stage weights as starting weights, the rim weighting was adjusted for all four measures, with the adjustment factor between the first and second stage weights trimmed at the 1st and 99th percentiles to dampen the extreme weights. The final weights were calculated as the first stage weights multiplied by the trimmed adjustment factor for the second stage, with confidence intervals for weighted prevalence estimates calculated using the “survey” package in R [30].

### Statistical Analyses

Statistical analyses were carried out in R [31]. To investigate the potential confounding effects of covariates on prevalence estimates we performed logistic regression on swab positivity as the outcome and: sex, age, region, employment type, ethnicity, household size and neighbourhood deprivation as explanatory variables. We adjusted for age and sex, and mutually adjusted for the other covariates to obtain odds ratio estimates and 95% confidence intervals.

We estimated adjusted vaccine effectiveness as 1 - odds ratio where the odds ratio was obtained from comparing vaccinated and unvaccinated individuals in a logistic regression model with swab positivity as outcome and with adjustment for age and sex, and age, sex, IMD quintile and ethnicity.

To estimate the underlying geographical variation in prevalence at local (sub-regional) level, we used a neighbourhood spatial smoothing method based on nearest neighbour up to 30 km. We calculated *N*_*n*_, the median number of study participants within 30 km of each study participant for each round or sub-round. We then calculated the local prevalence for 15 members of each LTLA as an estimate of the smoothed neighbourhood prevalence in that area.

To analyse trends in swab positivity over time, we used an exponential model of growth or decay with the assumption that the number of positive samples (from the total number of samples) each day arose from a binomial distribution. The model is of the form *I(t)* = *I*_0_ *e*^*rt*^., where *I(t)* is the swab positivity at time t, *I*_0_ is the swab positivity on the first day of data collection per round and r is the growth rate. The binomial likelihood for *P* (out of *N*) positive tests on a given day is then *P* ∼ *B*(*N, I*_0_. *e*^*rt*^.,) based on day of swabbing or, if unavailable, day of sample collection. We used a bivariate No-U-Turn sampler to estimate posterior credible intervals assuming uniform prior distributions on *I*_0_ and r [32]. We estimated the reproduction number R assuming a generation time that follows a gamma distribution with a shape parameter, n, of 2.29 and a rate parameter, *β*, of 0.36 (corresponding to a mean generation time of 6.29 days) [33]. R was estimated from the equation *R*=(1+ *r*/*β*)^*n* [34] using data from two sequential rounds and separately per round. We carried out a range of sensitivity analyses including estimation of R for different thresholds of Ct values that determine swab-positivity and for non-symptomatic individuals (not reporting symptoms on the day of swab or month prior).

We fit a Bayesian penalised-spline (P-spline) model [35] to the daily data using a No U-Turns Sampler in logit space, segmenting the data into approximately 5 day sections by regularly spaced knots, with further knots beyond the study period to minimise edge effects. We defined 4th order basis-splines (b-splines) over the knots with the final model consisting of a linear combination of these b-splines. We guarded against overfitting by including a second-order random-walk prior distribution on the coefficients of the b-splines, taking the form *b*_*i*_ = 2*b*_*i*−1_ – *b*_*i*−2_ + *u*_*i*′_, where *b*_*i*_ is the *ith* b-spline coefficient and *u*_*i*_ is normally distributed with *u*_*i*_ ∼ *N*(0, ϱ^2^) We assume a constant first derivative for the prior distribution which penalises against changes in the growth rate unless supported by the data as determined by the parameter ϱ for which we assume an inverse gamma prior distribution, ρ ∼ *IG*(0. 001, 0. 001). We assume the first two b-spline coefficients have uniform distribution, that is *b*_1_ and *b*_2_ are constant.

We compared daily prevalence data from rounds 1-13 of REACT-1 with publicly available national daily hospital admissions and COVID-19 mortality data (deaths within 28 days of a positive test). To do this we fit P-spline models as before to the daily hospital admissions and to the daily death data in order to obtain estimates for the expected number of outcomes on a given day. We then fit a simple two parameter model consisting of a lag time between the posterior of the P-spline estimate for each of hospitalisations or deaths, and the daily weighted prevalence calculated from REACT-1 data, and a scaling parameter, corresponding to the percentage of people who were swab-positive in the population on a particular day in comparison with future hospitalisations or deaths. Due to the time delay between the REACT-1 prevalence signal and daily hospitalisations and deaths the model was only fit to rounds 1-12. We then compared round 13 data to the estimated trend in hospitalisations and deaths to visualize any alterations in the link between these parameters and infection prevalence as measured in REACT-1. We estimated these relationships separately for: all ages, those aged under 65 years, and those 65 years and above.

To visualize the trends of the REACT-1 data over time we also fitted P-splines to all subsets of the REACT-1 data examined. For the REACT-1 data split by age (below 65 years and 65 years and above) we fit a mixed P-spline model in which a P-spline was fit separately to each age group but the smoothing parameter, ρ, was fit to both datasets simultaneously.

Further changes in the first derivative were assumed to happen at the same time for both datasets, with the condition *u*_*i*, <65_ – *u*_*i*, 65 +_ ∼ *N*(0, η^2^) and η given an uninformative prior distribution, η ∼ *IG* (0. 001, 0. 001).

### Viral genome sequencing

RT-PCR positive swab samples where there was sufficient sample volume and with N gene Ct values < 32 were sent frozen from the laboratory to the Quadram Institute, Norwich, UK for viral genome sequencing. Amplification of viral RNA used the ARTIC protocol [36] and sequencing libraries were prepared using CoronaHiT [37]. Analysis of sequencing data used the ARTIC bioinformatic pipeline [38] with lineages assigned using PangoLEARN [39].

We fit a Bayesian logistic regression model to the proportion of lineages that were identified as the Delta variant from round 10 to round 13 to obtain a daily growth rate advantage between Delta and other circulating lineages, Δ*r*. Assuming an exponential generation time of mean 6.29 days [33], the reproduction number, R, is given by 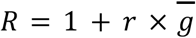[34]. The estimate of growth rate advantage can thus be converted into an additive R advantage through the equation 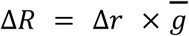, assuming the mean generation time is the same for all lineages. As a sensitivity the model was also fit to data from only round 11 to round 12 to check that edge effects were not biasing the calculation. The upper bound of prevalence for non-Delta lineages (none of which were detected in round 13) was estimated by calculating the 95% Wilson upper bound on the proportion of non-Delta lineage detected, then multiplying by the weighted prevalence estimate for round 13. This was then multiplied by the population of England to get an estimate for the upper bound on the average number of people infected with a non-Delta lineage at any one time during round 13.

### Public involvement

A Public Advisory Panel provides input into the design, conduct and dissemination of the REACT research programme.

### Ethics

We obtained research ethics approval from the South Central-Berkshire B Research Ethics Committee (IRAS ID: 283787).

## Data Availability

The datasets generated or analysed, or both, during this study are not publicly available because of governance restrictions.

## Data availability

Supporting data for tables and figures are available either: in this [spreadsheet](https://docs.google.com/spreadsheets/d/1HmGsdgNoJQHHSsRPyJBqU7nU0b58k-LOqJbSruLDIHs/edit#gid=1800680684); or in the inst/extdata directory of this [GitHub R package](https://github.com/mrc-ide/reactidd).

## Declaration of interests

We declare no competing interests.

## Funding

The study was funded by the Department of Health and Social Care in England. Sequencing was provided through funding from the COVID-19 Genomics UK (COG-UK) Consortium.

## Acknowledgements

SR, CAD acknowledge support: MRC Centre for Global Infectious Disease Analysis, National Institute for Health Research (NIHR) Health Protection Research Unit (HPRU), Wellcome Trust (200861/Z/16/Z, 200187/Z/15/Z), and Centres for Disease Control and Prevention (US, U01CK0005-01-02). GC is supported by an NIHR Professorship. HW acknowledges support from an NIHR Senior Investigator Award and the Wellcome Trust (205456/Z/16/Z). PE is Director of the Medical Research Council (MRC) Centre for Environment and Health (MR/L01341X/1, MR/S019669/1). PE acknowledges support from Health Data Research UK (HDR UK); the NIHR Imperial Biomedical Research Centre; NIHR HPRUs in Chemical and Radiation Threats and Hazards, and Environmental Exposures and Health; the British Heart Foundation Centre for Research Excellence at Imperial College London (RE/18/4/34215); and the UK Dementia Research Institute at Imperial (MC_PC_17114). We thank The Huo Family Foundation for their support of our work on COVID-19.

We thank key collaborators on this work – Ipsos MORI: Kelly Beaver, Sam Clemens, Gary Welch, Nicholas Gilby, Kelly Ward, Galini Pantelidou and Kevin Pickering; Institute of Global Health Innovation at Imperial College: Gianluca Fontana, Sutha Satkunarajah, Didi Thompson and Lenny Naar; North West London Pathology and Public Health England for help in calibration of the laboratory analyses; Patient Experience Research Centre at Imperial College and the REACT Public Advisory Panel; Quadram Institute, Norwich, UK: Thanh Le Viet, Nabil-Fareed Alikhan, Leigh M Jackson, Catherine Ludden; NHS Digital for access to the NHS register; the Department of Health and Social Care for logistic support; and the COVID-19 Taskforce of the Royal Statistical Society (UK) for helpful comments. SR acknowledges helpful discussion with attendees of meetings of the UK Government Scientific Pandemic Influenza – Modelling (SPI-M) committee.

Quadram authors gratefully acknowledge the support of the Biotechnology and Biological Sciences Research Council (BBSRC); their research was funded by the BBSRC Institute Strategic Programme Microbes in the Food Chain BB/R012504/1 and its constituent project BBS/E/F/000PR10352. We thank members of the COVID-19 Genomics Consortium UK (COG-UK) for their contributions to generating the genomic data used in this study. COG-UK is supported by funding from the MRC, part of UK Research & Innovation (UKRI), NIHR and Genome Research Limited, operating as the Wellcome Sanger Institute.

## Additional information

Full list of COG-UK author’s names and affiliations are available [here](https://drive.google.com/file/d/1KNedstJxnluNNa3MA2twbeTbuW6H11_X/view)

## Notes

\# Full list of consortium names and affiliations is provided as a supporting document

### Competing Interest Statement

The authors have declared no competing interest.

## References

1. Folegatti PM, Ewer KJ, Aley PK, Angus B, Becker S, Belij-Rammerstorfer S, et al. Safety and immunogenicity of the ChAdOx1 nCoV-19 vaccine against SARS-CoV-2: a preliminary report of a phase 1/2, single-blind, randomised controlled trial. Lancet. 2020;396: 467–478.

2. Polack FP, Thomas SJ, Kitchin N, Absalon J, Gurtman A, Lockhart S, et al. Safety and Efficacy of the BNT162b2 mRNA Covid-19 Vaccine. N Engl J Med. 2020. doi:10.1056/NEJMoa2034577

3. Coronavirus Pandemic (COVID-19). In: Our World in Data [Internet]. [cited 6 Apr 2021]. Available: https://ourworldindata.org/

4. Dhar MS, Marwal R, Vs R, Ponnusamy K, Jolly B, Bhoyar RC, et al. Genomic characterization and Epidemiology of an emerging SARS-CoV-2 variant in Delhi, India. bioRxiv. medRxiv; 2021. doi:10.1101/2021.06.02.21258076

5. Coronavirus (COVID-19) Vaccinations. In: Our World In Data [Internet]. [cited 21 Jul 2021]. Available: https://ourworldindata.org/covid-vaccinations

6. UK Government. UK government Covid-19 dashboard. In: UK government Covid-19 dashboard [Internet]. Available: https://coronavirus.data.gov.uk/

7. UK Government. Prime Minister sets out roadmap to cautiously ease lockdown restrictions. In: GOV.UK [Internet]. [cited 3 Feb 2021]. Available: https://www.gov.uk/government/news/prime-minister-sets-out-roadmap-to-cautiously-ease-lockdown-restrictions

8. Moving to step 4 of the roadmap. In: GOV UK [Internet]. 19 Jul 2021 [cited 21 Jul 2021]. Available: https://www.gov.uk/government/publications/covid-19-response-summer-2021-roadmap/moving-to-step-4-of-the-roadmap

9. Riley S, Atchison C, Ashby D, Donnelly CA, Barclay W, Cooke G, et al. REal-time Assessment of Community Transmission (REACT) of SARS-CoV-2 virus: Study protocol. Wellcome Open Research. 2020. p. 200. doi:10.12688/wellcomeopenres.16228.1

10. Riley S, Ainslie KEC, Eales O, Walters CE, Wang H, Atchison C, et al. Resurgence of SARS-CoV-2: Detection by community viral surveillance. Science. 2021;372: 990–995.

11. Office for National Statistics, UK. Coronavirus (COVID-19) Infection Survey, UK: 23 July 2021. 2021 Jul. Available: https://www.ons.gov.uk/peoplepopulationandcommunity/healthandsocialcare/conditionsanddiseases/bulletins/coronaviruscovid19infectionsurveypilot/23july2021

12. Office for National Statistics, UK. Population estimates for the UK, England and Wales, Scotland and Northern Ireland mid-2020. 2021 Jun. Available: https://www.ons.gov.uk/peoplepopulationandcommunity/populationandmigration/populationestimates/bulletins/annualmidyearpopulationestimates/mid2020

13. Wallinga J, van Boven M, Lipsitch M. Optimizing infectious disease interventions during an emerging epidemic. Proc Natl Acad Sci U S A. 2010;107: 923–928.

14. McLennan D, Noble S, Noble M, Plunkett E, Wright G GN. The English Indices of Deprivation 2019. 2019 Jan. Available: https://assets.publishing.service.gov.uk/government/uploads/system/uploads/attachment_data/file/833951/IoD2019_Technical_Report.pdf

15. Pritchard E, Matthews PC, Stoesser N, Eyre DW, Gethings O, Vihta K-D, et al. Impact of vaccination on new SARS-CoV-2 infections in the United Kingdom. Nat Med. 2021. doi:10.1038/s41591-021-01410-w

16. Public Health Scotland. Public Health Scotland COVID-19 Statistical Report as at 28 June 2021. 2021 Jun. Available: https://www.google.com/url?q=https://www.publichealthscotland.scot/media/8268/21-06-30-covid19-publication_report.pdf&sa=D&source=editors&ust=1627937430378000&usg=AOvVaw2Kwz_u0_KQraqrxqTW-xyX

17. Lopez Bernal J, Andrews N, Gower C, Gallagher E, Simmons R, Thelwall S, et al. Effectiveness of Covid-19 Vaccines against the B.1.617.2 (Delta) Variant. N Engl J Med. 2021. doi:10.1056/NEJMoa2108891

18. Dagan N, Barda N, Kepten E, Miron O, Perchik S, Katz MA, et al. BNT162b2 mRNA Covid-19 Vaccine in a Nationwide Mass Vaccination Setting. N Engl J Med. 2021. doi:10.1056/NEJMoa2101765

19. Ministry of Health, Israel. Vaccine efficacy among those first vaccinated. In: GOV IL [Internet]. 18 Jul 2021 [cited 8 Mar 2021]. Available: https://www.gov.il/BlobFolder/reports/vaccine-efficacy-safety-follow-up-committee/he/files_publications_corona_two-dose-vaccination-data.pdf

20. Saxena S, Skirrow H, Wighton K. Should the UK vaccinate children and adolescents against covid-19? BMJ. 2021;374: n1866.

21. Planas D, Veyer D, Baidaliuk A, Staropoli I, Guivel-Benhassine F, Rajah MM, et al. Reduced sensitivity of SARS-CoV-2 variant Delta to antibody neutralization. Nature. 2021. doi:10.1038/s41586-021-03777-9

22. Khoury DS, Cromer D, Reynaldi A, Schlub TE, Wheatley AK, Juno JA, et al. Neutralizing antibody levels are highly predictive of immune protection from symptomatic SARS-CoV-2 infection. Nat Med. 2021;27: 1205–1211.

23. Haas EJ, Angulo FJ, McLaughlin JM, Anis E, Singer SR, Khan F, et al. Impact and effectiveness of mRNA BNT162b2 vaccine against SARS-CoV-2 infections and COVID-19 cases, hospitalisations, and deaths following a nationwide vaccination campaign in Israel: an observational study using national surveillance data. Lancet. 2021. doi:10.1016/S0140-6736(21)00947-8

24. Stowe J, Andrews N, Gower C, Gallagher E, Utsi, L, Simmons, R. Effectiveness of COVID-19 vaccines against hospital admission with the Delta (B.1.617.2) variant. Public Health England Library. Public Health England; Available: https://khub.net/web/phe-national/public-library/-/document_library/v2WsRK3ZlEig/view_file/479607329?_com_liferay_document_library_web_portlet_DLPortlet_INSTANCE_v2WsRK3ZlEig_redirect=https%3A%2F%2Fkhub.net%3A443%2Fweb%2Fphe-national%2Fpublic-library%2F-%2Fdocument_library%2Fv2WsRK3ZlEig%2Fview%2F479607266

25. Sheikh A, McMenamin J, Taylor B, Robertson C, Public Health Scotland and the EAVE II Collaborators. SARS-CoV-2 Delta VOC in Scotland: demographics, risk of hospital admission, and vaccine effectiveness. Lancet. 2021;397: 2461–2462.

26. Thomas SJ, Moreira ED Jr, Kitchin N, Absalon J, Gurtman A, Lockhart S, et al. Six month safety and efficacy of the BNT162b2 mRNA COVID-19 vaccine. bioRxiv. medRxiv; 2021. doi:10.1101/2021.07.28.21261159

27. REal-time Assessment of Community Transmission (REACT) study. In: Imperial College [Internet]. Available: https://www.imperial.ac.uk/medicine/research-and-impact/groups/react-study/

28. Sharot T. Weighting survey results. 1986 [cited 12 Jan 2021]. Available: http://redresearch.com/wp/wp-content/uploads/2016/01/Weighting-Survey-Results.pdf

29. Park N. Population estimates for the UK, England and Wales, Scotland and Northern Ireland - Office for National Statistics. Office for National Statistics; 23 Jun 2020 [cited 6 Mar 2021]. Available: https://www.ons.gov.uk/peoplepopulationandcommunity/populationandmigration/populationestimates/bulletins/annualmidyearpopulationestimates/latest

30. Lumley T. Analysis of Complex Survey Samples. Journal of Statistical Software, Articles. 2004;9: 1–19.

31. R Core Team. R: A Language and Environment for Statistical Computing. R Foundation for Statistical Computing; 2020. Available: https://www.R-project.org/

32. Hoffman MD, Gelman A. The No-U-Turn Sampler: Adaptively Setting Path Lengths in Hamiltonian Monte Carlo. arXiv [stat.CO]. 2011. Available: http://arxiv.org/abs/1111.4246

33. Bi Q, Wu Y, Mei S, Ye C, Zou X, Zhang Z, et al. Epidemiology and transmission of COVID-19 in 391 cases and 1286 of their close contacts in Shenzhen, China: a retrospective cohort study. Lancet Infect Dis. 2020;20: 911–919.

34. Wallinga J, Lipsitch M. How generation intervals shape the relationship between growth rates and reproductive numbers. Proceedings Of The Royal Society B-Biological Sciences. 2006;274: 599–604.

35. Lang S, Brezger A. Bayesian P-Splines. J Comput Graph Stat. 2004;13: 183–212.

36. Quick J. nCoV-2019 sequencing protocol v3 (LoCost). 2020 [cited 4 May 2021]. Available: https://www.protocols.io/view/ncov-2019-sequencing-protocol-v3-locost-bh42j8ye

37. Baker DJ, Aydin A, Le-Viet T, Kay GL, Rudder S, de Oliveira Martins L, et al. CoronaHiT: high-throughput sequencing of SARS-CoV-2 genomes. Genome Med. 2021;13: 21.

38. A Nextflow pipeline for running the ARTIC network’s field bioinformatics tools. Github; Available: https://github.com/connor-lab/ncov2019-artic-nf

39. Phylogenetic Assignment of Named Global Outbreak LINeages (PANGOLIN). Github; Available: https://github.com/cov-lineages/pangolin

